# Pre-vaccination carriage prevalence of *Streptococcus pneumoniae* serotypes among internally displaced people in Somaliland

**DOI:** 10.1101/2024.02.09.24302568

**Authors:** Kevin van Zandvoort, Abdirahman Ibrahim Hassan, Mohamed Bobe, Casey L. Pell, Mohammed Saed Ahmed, Belinda D. Ortika, Saed Ibrahim, Mohamed Ismail Abdi, Mustapha A Karim, Rosalind M Eggo, Sulieman Yusuf, Jason Hinds, Saed Mohamood Soleman, Rachael Cummings, Catherine McGowan, Kim Mulholland, Mohamed Abdi Hergeeye, Catherine Satzke, Francesco Checchi, Stefan Flasche

**Affiliations:** Department of Infectious Disease Epidemiology, London School of Hygiene & Tropical Medicine. United Kingdom.; Republic of Somaliland Ministry of Health Development. Somaliland; Save the Children International Somaliland. Somaliland; Infection, Immunity and Global Health, Murdoch Children’s Research Institute, Melbourne, Australia; Institute for Infection and Immunity, St. George’s, University of London, United Kingdom; BUGS Bioscience, London Bioscience Innovation Centre, London, United Kingdom; Save the Children UK. United Kingdom; Department of Microbiology and Immunology at the Peter Doherty Institute for Infection and Immunity, The University of Melbourne, Melbourne, Australia; Department of Paediatrics, The University of Melbourne, Melbourne, Australia

## Abstract

Populations affected by humanitarian crises likely experience high burdens of pneumococcal disease. Streptococcus pneumoniae carriage estimates are essential to understand pneumococcal transmission dynamics and the potential impact of pneumococcal conjugate vaccines (PCV). Over 100 million people are forcibly displaced worldwide, yet here we present only the second pneumococcal carriage estimates for a displaced population.

In October 2019, we conducted a cross-sectional survey among internally displaced people (IDP) living in Digaale, a permanent IDP camp in Somaliland where PCV has not been implemented. We collected nasopharyngeal swab samples from 453 residents which were assessed for presence of pneumococci and serotyped using DNA microarray.

We found that pneumococcal carriage prevalence was 36% (95%CI 31 – 40) in all ages, and 70% (95%CI 64 – 76) in children under 5. The three most common serotypes were vaccine serotypes 6B, 19F, and 23. We estimated that the serotypes included in the 10-valent PNEUMOSIL vaccine were carried by 41% (95%CI 33 – 49) of all pneumococcal carriers and extrapolated that they caused 52% (95%CI 35 – 72) of invasive pneumococcal disease. We found some evidence that pneumococcal carriage was associated with recent respiratory symptoms, the total number of physical contacts made, and with malnutrition in children under 5. Through linking with a nested contact survey we projected that pneumococcal exposure of children under 2 was predominantly due to contact with children aged 2-5 (39%; 95%CI 32 – 48) and 6-14 (25%; 95%CI 18 – 33).

These findings suggest considerable potential for direct and indirect protection against pneumococcal disease in Digaale through PCV use in children and potentially adolescents.

## Introduction

The United Nations High Commissioner for Refugees estimates that over 100 million people were forcibly displaced worldwide in 2022, of whom over half are internally displaced and more than 40% are children (1). These people typically live in overcrowded settings, have poor access to hygiene and healthcare, and thus experience high morbidity and mortality from respiratory diseases (2–4) including invasive disease from *Streptococcus pneumoniae* (pneumococcus) (5). There is a lack of health research in these populations in general (6), and for pneumococcal disease specifically (2,4). Pneumococcal prevalence estimates are only available for one other displaced population globally, living in Mae La refugee camp, Thailand (7).

Pneumococcal Conjugate Vaccines (PCV) are highly efficacious and have been used routinely for protecting children against pneumococcal colonization and disease in most countries worldwide. Five PCVs are currently used for childhood immunization: Prevenar 20 (20 valent, targeting 20 of the over 100 serotypes), Vaxneuvance (15 valent), Prevenar 13 (13 valent), Synflorix and PNEUMOSIL (both 10 valent) (8–10). However, despite the high prevalence of risk factors for severe disease and intense transmission, they are rarely offered to populations that have become displaced as a result of food insecurity, conflict, natural disasters, or other emergencies (2).

Pneumococcal colonization is common and is a precursor to disease, thus providing an opportunity to assess the likely risk for disease, even in small populations, without costly disease surveillance programmes that would be difficult to establish in displaced populations (11). Such information is crucial to inform the design of effective pneumococcal immunization strategies in displaced populations where routine immunization is rarely possible (2). We conducted a cross-sectional survey to estimate nasopharyngeal carriage prevalence and related risk factors in Digaale, a camp for internally displaced people (IDPs) in Somaliland (2). PCVs had not yet been introduced in Somaliland at the time of writing.

## Methods

### Study population and sampling method

We conducted a cross-sectional survey in October-November 2019. The Digaale IDP camp was established in 2014 and, at the time of our study, housed an estimated population of 3,000 people largely displaced due to drought and food insecurity during 2013 and 2014 (12). The camp is located about 4km south-east of Hargeisa, the capital of Somaliland, and consists of corrugated-steel shelters. The population is served by a school and primary health centre.

We visited all 894 shelters in Digaale and invited households to participate in our survey. We first administered a structured household survey among consenting households to establish their composition and collect household-level information on shelter conditions, pneumococcal risk factors, and retrospective mortality and demographic changes. We then selected and invited individual household members for participation in a survey on carriage, contact and individual-level risk factors. We aimed to sample 100 individuals in each of the following age groups: <1, 1, 2-5, 6-14, 15-29, 30-49, and ≥50 years (y) old to detect age-specific pneumococcal prevalence within 10% precision. We purposively oversampled young children who have the highest incidence of pneumococcal carriage and disease. Quota sampling by age was used to select individual household members.

We returned to individual participants two days after the household survey and study enrolment to conduct the contact and risk factor survey, in which we asked about individual-level risk factors including social contacts, and measured anthropometry for children aged 6-59 months. Further details about the design and sampling method, as well as detailed social contact and household-level findings, are described in Van Zandvoort et al (12). In the final week of data collection, one to four weeks after participation in the contact survey, we followed up participants to collect a nasopharyngeal swab sample. To compensate for loss-to-follow up, additional participants were sampled from household members of participating households.

All swabs were collected at a community hall in the centre of the camp. During swab collection, we asked participants whether they had experienced any respiratory symptoms (cough, sore throat, sneezing, wheezing, headache, or fever) or used any antibiotics in the two weeks prior. Responses for children under 10y were provided by an adult parent or caregiver. Participants with a contraindication for a nasopharyngeal swab such as facial trauma were excluded.

### Sample collection, storage, and shipment

Trained nurses collected nasopharyngeal swabs from each participant using flexible paediatric-(Ultra Minitip) or adult-size (Flexible Minitip) flocked swabs (FLOQSwabs; Copan Diagnostics, USA), following WHO recommendations (13,14). Paediatric-sized swabs were used for children under 15y. The nasopharyngeal swabs were stored in screw-capped tubes containing 1 ml of skim milk-tryptone-glucose-glycerol (STGG) medium and kept on wet ice in cool boxes immediately after collection. Samples were transferred to a −20°C freezer at the Ministry of Health Development national cold chain facility within eight hours of collection (15). Within two weeks after collection, all samples were transferred to an ultra-low temperature (ULT) freezer at the culture laboratory of the Somaliland National Tuberculosis Hospital. Due to technical issues with the ULT freezer, samples were temporarily stored at −20°C for 15 days in March 2020.

We used a prequalified shipping solution with phase change materials (PCMs) that provided passive cooling to keep contents below −15°C for up to 96h as per manufacturer specifications (Schaumaplast, DE) (16). We found that it maintained temperatures below −15°C up to 160h in an empty trial shipment from London to Hargeisa when conditioned at ULT (Supplemental Material Section B). Samples were first shipped to Nairobi, Kenya, where they were repackaged and placed on dry ice for further transport to the Murdoch Children’s Research Institute (MCRI) in Melbourne, Australia. We successfully completed a pilot shipment of 81 samples in May 2021, and shipped all remaining samples in December 2021. Transit delays during the second shipment extended the period during which only passive cooling was provided by PCMs to 11 days. There was no temperature monitoring during this second shipment, but we project that temperatures may have increased to >0°C for up to 2.5 days based on temperature measurements from the pilot shipment. We explored any difference in carriage estimates between the two shipments in a sensitivity analysis.

### Microbiological analysis

Upon arrival at the MCRI laboratory, STGG swab samples were immediately stored at ULT until testing. Briefly, samples were thawed, vortexed, and DNA extracted from 100 µl using the QIAcube HT machine (Qiagen) following a protocol described previously (17). Concurrently, each sample was cultured overnight on selective agar, and growth harvested if alpha-haemolytic colonies were present (18). Real-time quantitative PCR, targeting the *lytA* gene (*lytA* qPCR) (19), was conducted as previously described, except for using 5 µl template DNA and the AriaMX PCR system (Agilent) (18). For samples with presumptive pneumococci (*lytA* qPCR cycle threshold <40 and alpha-haemolytic growth), DNA was extracted from harvested colonies using the QIAcube HT machine (Qiagen) and the resultant DNA serotyped using microarray (Senti-SP version 1.5, BUGS Bioscience) (18). Pneumococcal density was calculated using a genomic DNA standard curve (18). Serotype-specific density was calculated by multiplying the pneumococcal density (as determined by *lytA* qPCR) by the relative abundance of the serotype (as determined by microarray). Carriage density estimates were log10-transformed and reported as log_10_ genome equivalents/ml (GE/ml).

### Statistical analysis

Serotypes were grouped as non-encapsulated (NESp), vaccine serotypes (VT) for each of PNEUMOSIL, Synflorix, Prevenar 13, Vaxneuvence, and Prevenar 20 vaccines, or non-vaccine serotypes (NVT). Unless stated otherwise, we used PNEUMOSIL targeted serotypes to define VT in our base case analysis, because PNEUMOSIL was specifically developed to provide a lower cost alternative that targets the main serotypes causing pneumococcal disease in low and middle income countries (20). Analyses that defined VT using the other vaccines are presented in the Supplementary Material.

We estimated the population and age-specific prevalence of pneumococcal serotypes with 95% binomial confidence intervals. To account for multiple serotype carriage, serotypes were weighted by their relative abundance within a sample to calculate the serotype distribution including co-carried serotypes, so that the weights of all serotypes in a sample summed to one. Unweighted distributions of all and dominant-only serotypes were assessed in a sensitivity analysis. We define serotypes ranked with the highest relative abundance in a single sample as dominant serotypes, and used logistic regression to assess the odds that serotypes were dominant.

As we sampled a large proportion of households (65%) and individuals (17%) living in Digaale, we used finite population corrections (FPC) to calculate standard errors used in population-level estimates. To correct for imbalance resulting from quota sampling and thus improve the representativeness, we applied poststratification weights based on age group and gender (12,21). We assessed the sensitivity of our population level estimates to poststratification in additional analyses. We used the *Survey* package in *R* to perform poststratification weighting and to apply the FPC when estimating weighted means, proportions, and quantiles where applicable (22).

To project the proportion of current IPD cases caused by serotypes covered by PCVs, we combined the dataset with age-and serotype-specific estimates of invasiveness by Løchen et al (23), and computed confidence intervals by bootstrapping our dataset and invasiveness estimates. More details are provided in the Supplemental Material.

To estimate the likely contribution of different age groups to pneumococcal exposure and transmission, we followed a method developed by Qian et al (24) and calculated the proportion of colonisations attributable to contact with different age groups by linking the estimates of carriage prevalence with previously reported contact patterns (12) in those individuals and the likelihood of colonisation among those contacts. As the contact and carriage datasets featured different statistical error processes, we computed confidence intervals for this analysis through bootstrapping from each parameter’s uncertainty distribution.

We used logistic regression to test the univariate association of likely risk factors with the odds of pneumococcal carriage and used linear regression to test their association with mean logged overall carriage density in pneumococcal carriers. Age and gender were included as *a priori* defined confounding variables in both analyses, but no other multivariate analyses were conducted due to data sparsity. We also used logistic regression to test for any association between the odds of multiple carriage and age. FPC and poststratification weights were not applied in regression analyses.

All analyses were conducted in *R4.2.2*. Analysis scripts and anonymised aggregated data will be made available on final publication.

### Ethical approval

Ethical approval for the study was granted by the Research Ethical Committee of the London School of Hygiene and Tropical Medicine (16577) and the Republic of Somaliland Ministry of Health Development (2/13075/2019). The funding sources had no role in the study design; collection, analysis, and interpretation of data; or in writing the report.

## Results

### Participant sampling and enrolment

We enrolled 464 (65%) households and collected demographic data from 2,049 individuals living in these households. A contact and individual-level risk-factor survey was conducted among 509 participants. We collected a nasopharyngeal swab from 365 of these participants. An additional 88 nasopharyngeal swabs were collected from other individuals from consenting households. In total, 453 swabs were collected (Figure 1).

**FIGURE 1.**
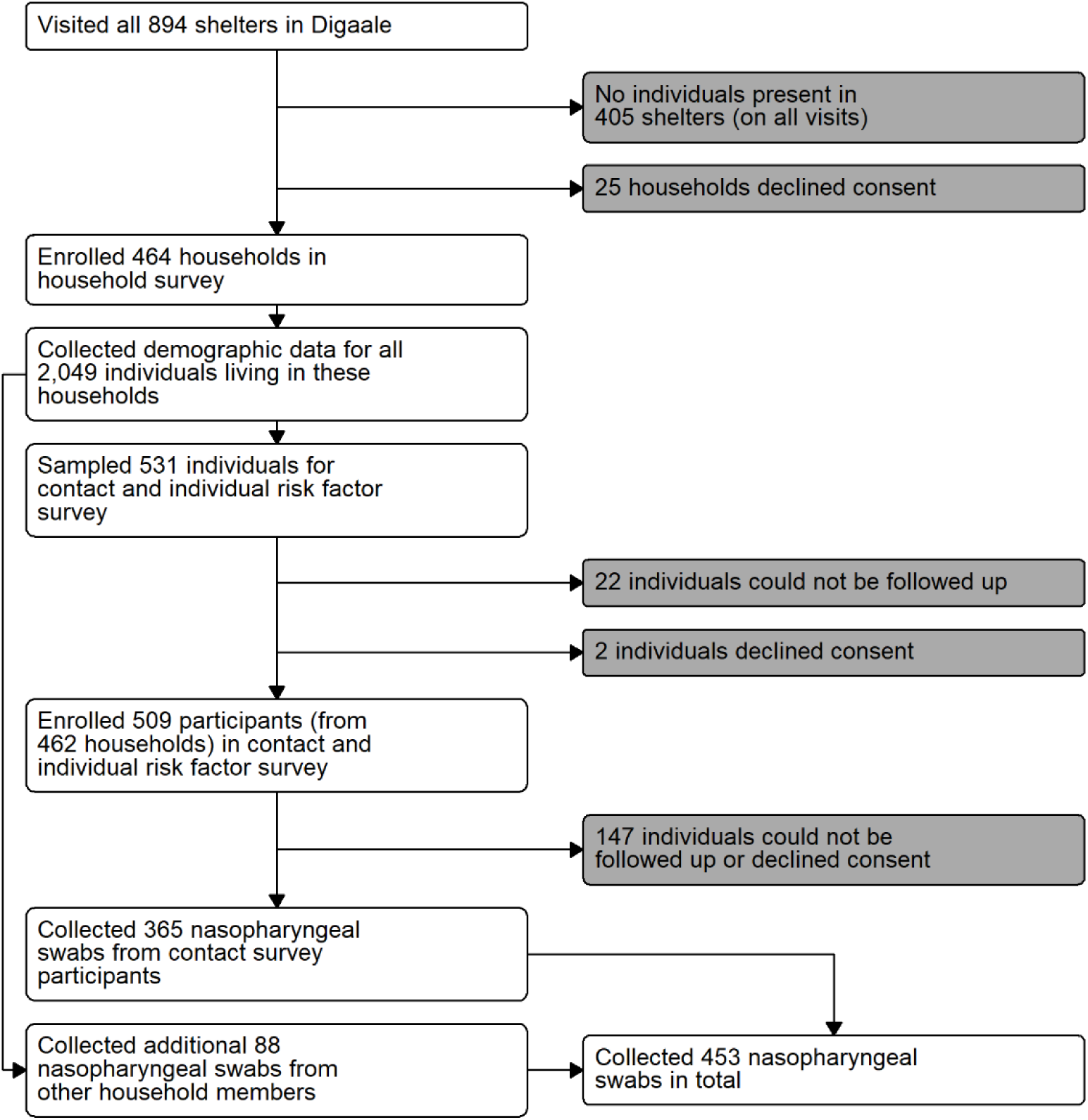
FLOWCHART OF SAMPLING PROCEDURE. All households that were present were invited to participate in the survey. A structured household survey was conducted in consenting households. Individual household members were selected using quota sampling and invited for a contact and individual-level risk factor survey. Participants were followed up after one to four weeks to administer a nasopharyngeal swab. Additional household members were sampled from consenting households using quota sampling to compensate for loss to follow-up.

### Sample characteristics

Two (0.4%) nasopharyngeal swab samples were excluded due to insufficient sample volume for laboratory testing. An additional two (0.4%) samples were *lytA* positive, indicating pneumococcal colonization, but with no growth prohibiting microarray serotyping; these were included only in non-serotype specific carriage prevalence analyses. Due to data entry errors, 53 swabs (12%) could not be fully linked to records in contact or household datasets. This included 16 swabs that could not be linked to their household information, and 6 samples without information on age and gender (Supplemental Material Section A). Data from these swabs are excluded in analyses that require linking to those data, but included otherwise.

67% of the study participants from whom swabs were collected were female (Table 1). Median age was 13y and median household size was 5 people. Pneumococci were detected in 39% (175/445) of samples, with at least one PNEUMOSIL VT present in 49% (85/175) of positive samples. We estimated the overall carriage prevalence in Digaale as 36% (95%CI 31 – 40%), with 41% (95%CI 33 – 49%) of carriers carrying at least one VT. Estimated prevalence was 70% (95%CI 64 – 76%) in children under 5y, with VTs carried by 61% (95%CI 53 – 69) of carriers. Large proportions of participants reported respiratory symptoms (64%) and antibiotic use (36%) in the two weeks leading up to the sampling.

**TABLE 1.**
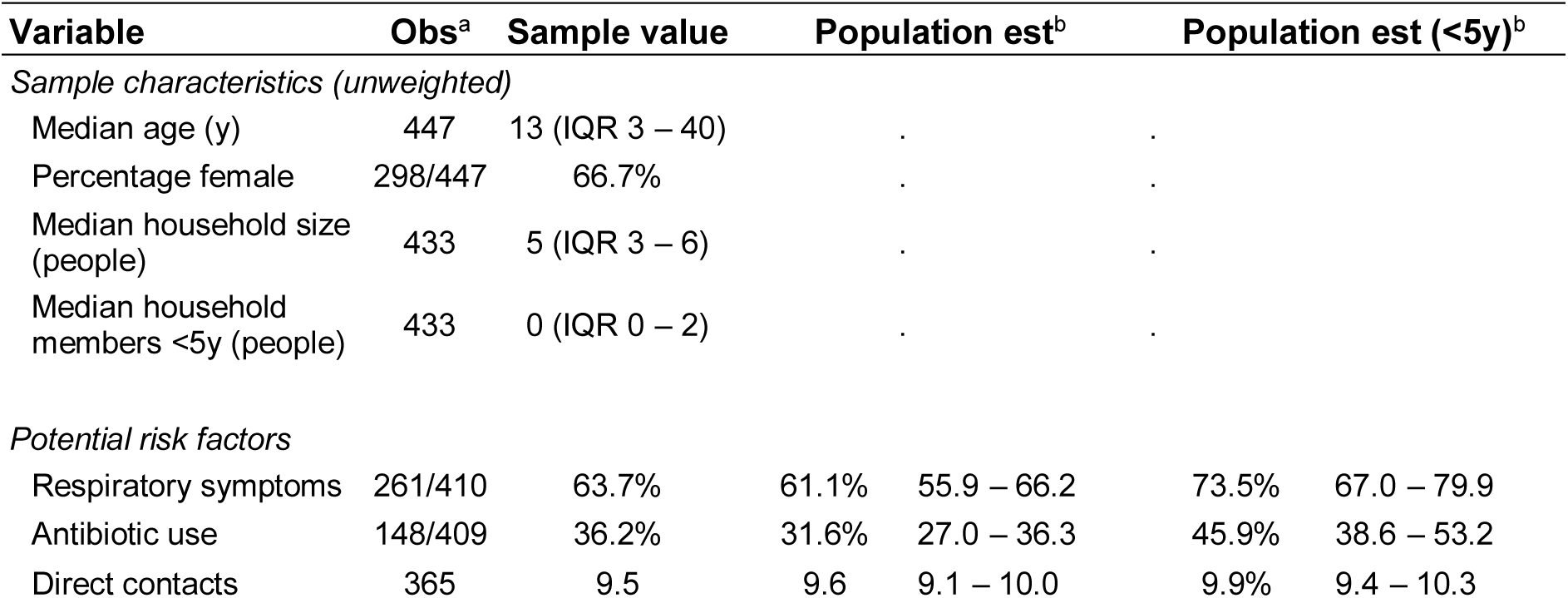
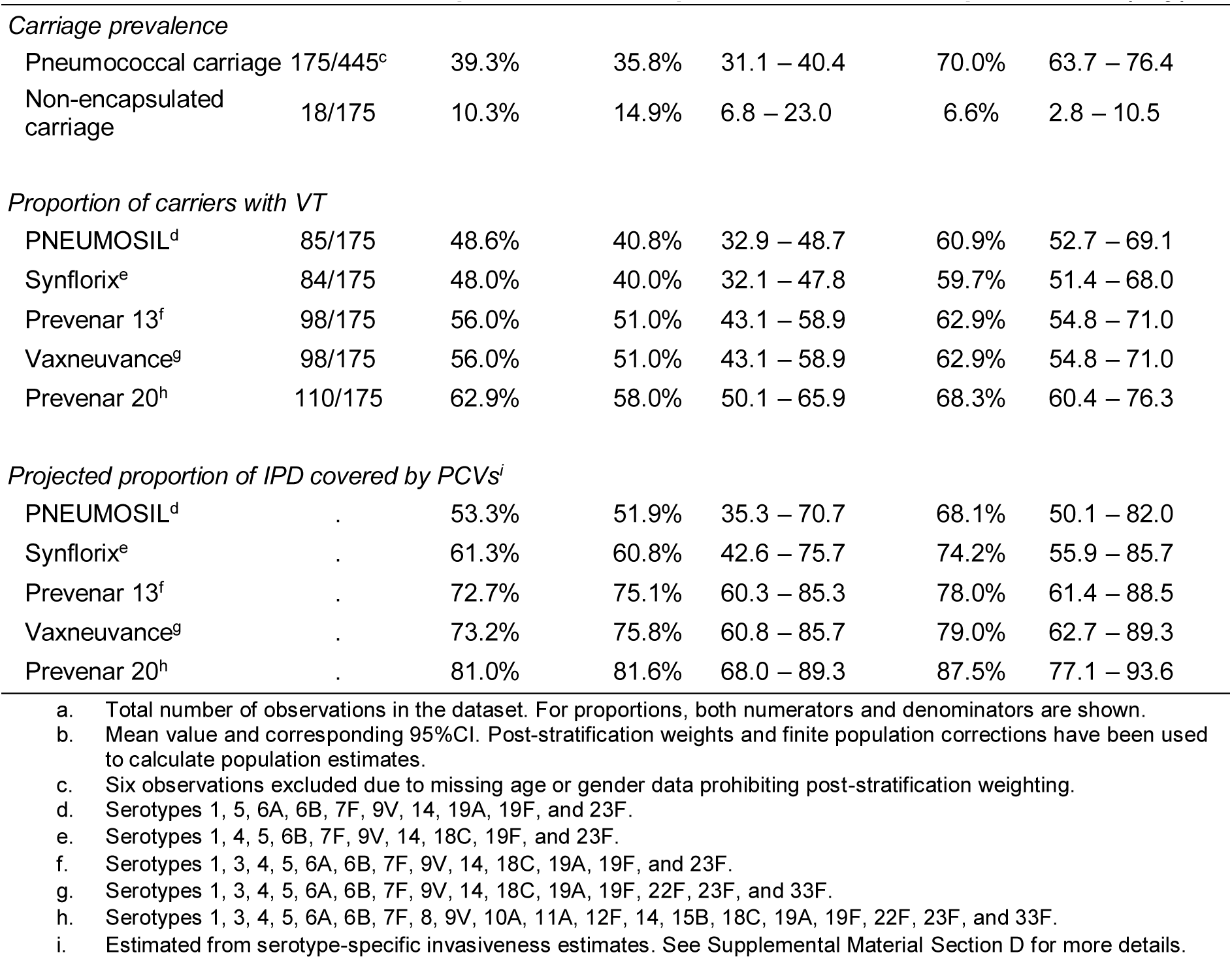
SAMPLE CHARACTERISTICS, CARRIAGE PREVALENCE, AND INVASIVE DISEASE LIKELY CAUSED BY VACCINE SEROTYPES.

### Serotype distribution

The three most prevalent pneumococcal serotypes overall (6B, 19F, and 23F) were VTs included in all PCVs, followed by three serotypes (6C, 11A, 16F) of which 11A is only included in Prevenar 20, and all others are NVTs (Figure 2). A similar serotype distribution was observed in children under 5y, while 15B/C, 6B, and 6C were the most prevalent serotypes in people over 5y (Supplemental Figure C2). There was little difference in the serotype distribution when restricting analysis to dominant serotypes alone, or without weighting for relative abundance in multiple serotype carriers (Supplemental Figure C1). We extrapolate that 52% (95%CI 35 – 71) of all IPD cases and 68% (95%CI 50 – 82) of IPD cases in <5y were caused by VT serotypes, with slightly increased proportions for higher valency vaccines (Table 1).

**FIGURE 2.**
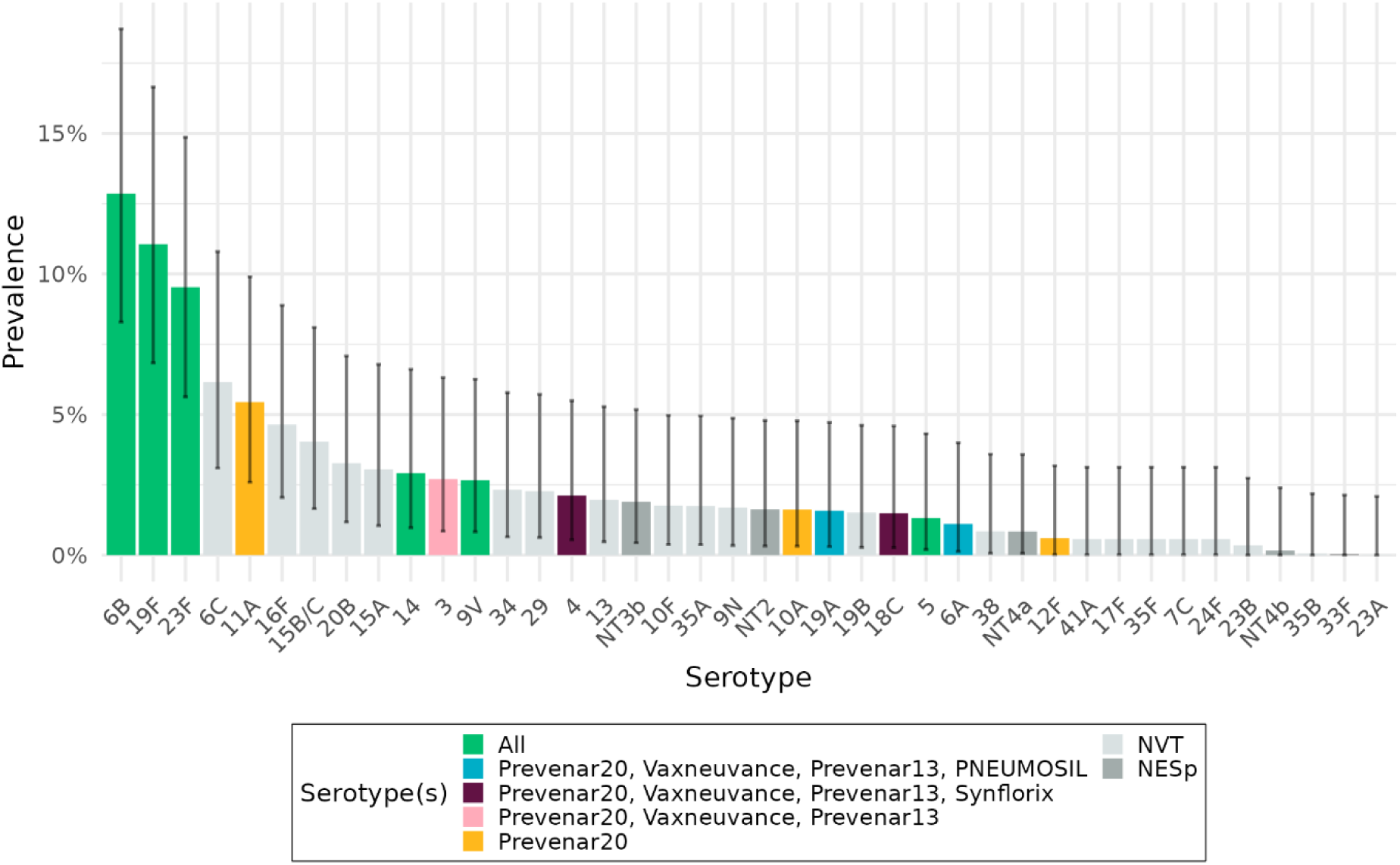
PNEUMOCOCCAL SEROTYPE DISTRIBUTION. Bars show the carriage prevalence of pneumococcal serotypes identified in Digaale IDP camp, weighted by their relative abundance. Coloured bars show the serotypes included in the five PCVs, dark grey bars non-encapsulated pneumococci, and light grey bars other serotypes not included in the PCVs. Error bars show 95% confidence intervals for each estimate.

### Multiple serotype carriage

Co-colonization with more than one serotype was detected in 30% (52/175) of samples with pneumococci, corresponding to a population-level prevalence of 39% (95%CI 35 – 44%). There were 36 samples in which two serotypes were detected, 16 samples with three serotypes, and three samples with more than three serotypes. The odds that the serotype was dominant was 2.0 (95%CI 1.1 – 3.7) times higher for VTs than for NVTs in all carriers, and 2.8 (95%CI 1.0 – 8.0) times higher among carriers colonized with both VT and NVT (Supplemental Table C2).

### Sensitivity analyses

We found no evidence of a difference in pneumococcal carriage, density of pneumococcal serotypes, number of carried serotypes, or serotype distribution between our two sample shipments (Supplemental Material Section B). Post stratifying our estimates did not substantially affect pneumococcal carriage prevalence estimates (Supplemental Table C5).

### Antimicrobial resistance

Microarray assays detected the presence of select antimicrobial resistance genes in 30% (95%CI 21 – 41%) of samples (Supplemental Table C2). In those samples, the most common detected genes typically associated with antimicrobial resistance were *tetM* (28% [95%CI 19 – 39]) and *ermB* (9% [95%CI 4 – 16]). We restricted this analysis to samples in which no other species and only a single pneumococcal serotype were detected

### Carriage prevalence and serotype distribution by age

Overall carriage prevalence was 79% (95%CI 72 – 87%) and 67% (95%CI 58 – 75%) in children under 2 and 2-5y, respectively. Carriage prevalence was 41% (95%CI 30 – 51%) in children aged 6-14y, 28% (95%CI 16 – 41%) in people aged 15-29y, 18% (95%CI 7 – 28%) in adults aged 30-49y and 8% (95%CI 4 – 12) in adults aged ≥50y (Figure 3). Co-colonization rates decreased by age alongside reductions in overall prevalence, although this reduction was not statistically significant. The proportion of VT among all carriers was similar across age groups and robust to the definition of VT for different PCV products (Supplemental Figure C3).

**FIGURE 3.**
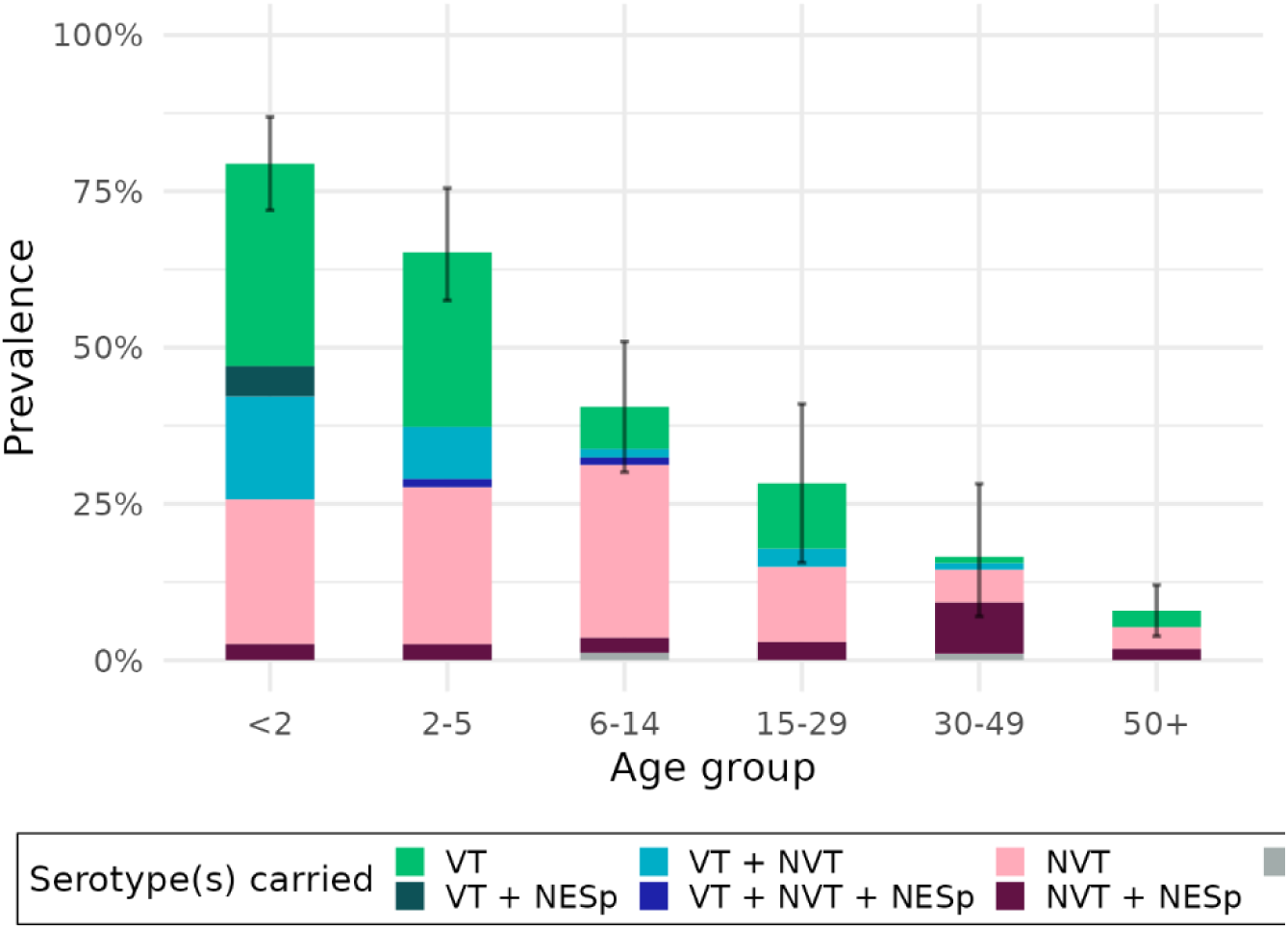
PREVALENCE AND SEROTYPE DISTRIBUTION BY AGE. Bars show the estimated prevalence of pneumococcal serotypes by age group, weighted for age and gender. Error bars show 95% confidence intervals around overall pneumococcal carriage prevalence. Colours show the prevalence of serotypes that are carried; VT: only vaccine type(s), NVT: only non-vaccine type(s); NT: only non-encapsulated type(s); VT + NVT: both vaccine-and non-vaccine type(s). Multiple carriage with non-encapsulated type(s) is shown as a darker shading.

### Contribution of different age groups to pneumococcal exposure

We projected that a large proportion (39% [95%CI 32 – 48]) of pneumococcal exposure of children <2y may be attributed to contact with 2-5y children, followed by school age children aged 6-14y (25% [95%CI 18 – 33]) (Figure 4 and Supplemental Table C6). A similar contribution was made by carriers of these age groups to exposure of children aged 2-5y (45% [95%CI 38 – 53]; and 30% [95%CI 22 – 38]). Most of the exposure of school age children, however, was found to be attributable to other school age children (51% [95%CI 42 – 60]), followed by 2-5y olds (26% [95%CI 20 – 32]). While carriage prevalence was high in children <2y, this age group was found to contribute relatively little to onward transmission to any age group.

**FIGURE 4.**
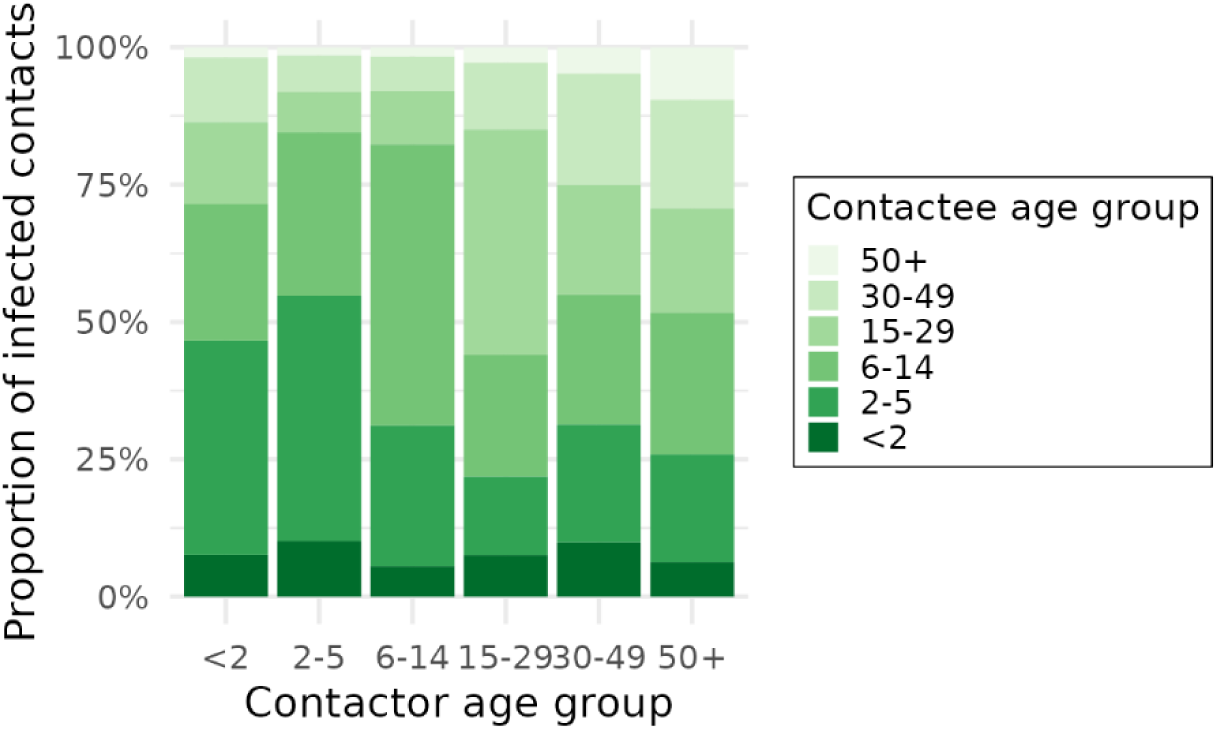
THE CONTRIBUTION OF DIFFERENT AGE GROUPS TOWARDS THE AGE-SPECIFIC EXPOSURE TO PNEUMOCOCCUS. Bars show the average proportion of contacts made by a contactor of age *i* (x-axis), with pneumococci-carrying contactees of different age groups. Shades of green stratify into age group of the contactee, i.e. the person potentially transmitting to the contactor.

### Association of risk factors with pneumococcal carriage and carriage density

We found no evidence that the number of overall household members increased the odds of pneumococcal carriage, but weak evidence that living with one additional household member <5y of age increased the odds of carriage by 1.3 (95%CI 1.0 – 1.8) (Table 2). The odds of carriage were 2.0 (95%CI 1.2 – 3.3) times higher in people with recent respiratory symptoms. Having a cough (2.0 [95%CI 1.2 – 3.3]) had the strongest association, followed by having a sore throat (1.7 [95%CI 1.0 – 3.0]). There was some evidence that the odds of carriage increased by 1.1 (95%CI 1.0 – 1.2) for every additional physical contact reported. We found good evidence for a reduction in the odds of carriage for improved scores of weight-for-age (0.6 [95%CI 0.4 – 0.9]) and height-for-age (0.6 [95%CI 0.4 – 0.8]) among children 6-59 months old, but no evidence for an association with weight-for-height or middle-upper arm circumference. Notably, we did not find any evidence of an association with self-reported antibiotic use.

**TABLE 2.**
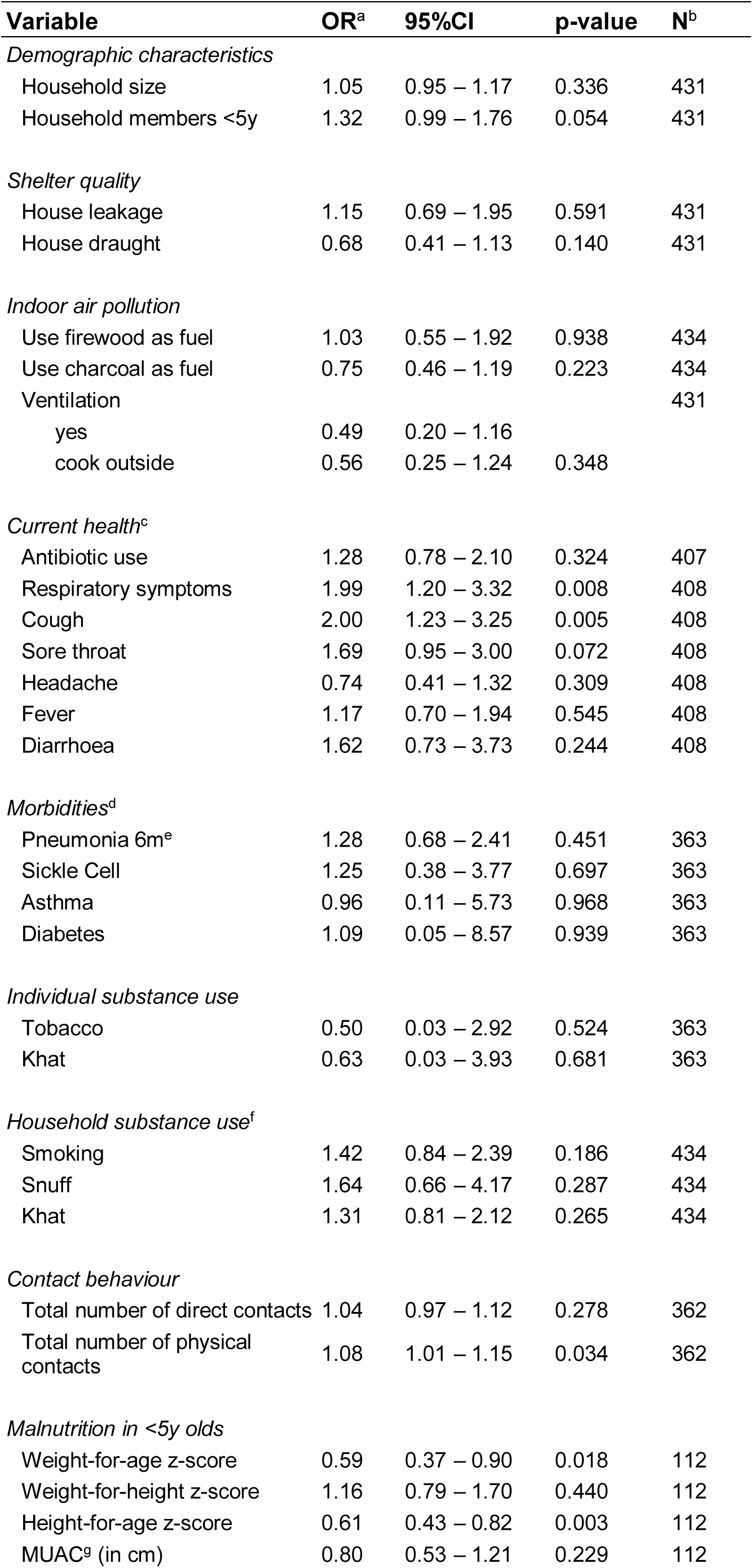
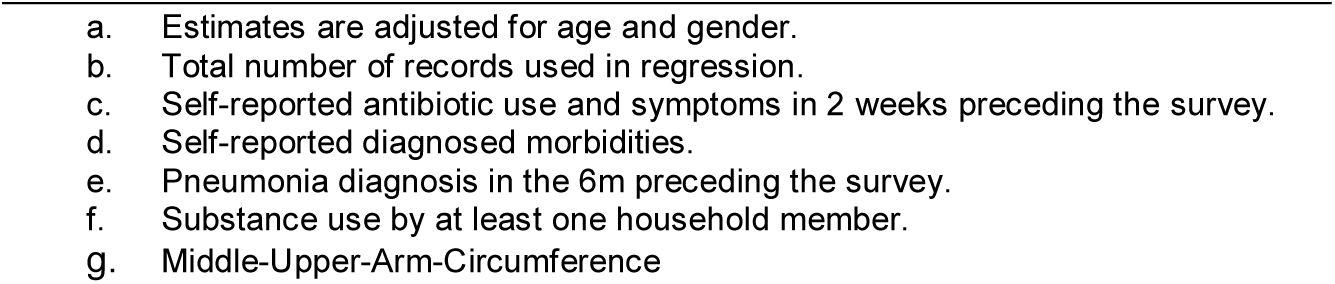
ASSOCIATION BETWEEN RISK FACTORS AND PNEUMOCOCCAL CARRIAGE.

We also tested the association between these risk factors and the density of pneumococcal carriage (Supplemental Table C3) and found weak evidence that living with one additional household member <5y was associated with a small 0.2 (95%CI −0.0 – 0.4) increase in mean log10 GE/ml, while associations with respiratory symptoms were either non-significant or negative: the mean log10 density was 0.40 (95%CI −0.8 – 0.0) lower in participants reporting a sore throat in the two weeks preceding the survey. Again, we did not find any significant association with self-reported antibiotic use. There was very weak evidence for an increase in children’s mean log10 density with a one-unit increase in weight-for-height z-score (0.2 [95%CI −0.0 – 0.4]).

## Discussion

This is the first study to have estimated pneumococcal serotype prevalence in Somaliland and in an IDP camp. We find high carriage prevalence of 36% in all age groups, and 70% in children under 5y. Between 40 and 58% of pneumococcal carriers carried serotypes included in PCVs, depending on the PCV product, and the three most prevalent serotypes were covered by all PCVs. The majority of exposure to pneumococcal carriers in children younger than 15y may have been attributable to carriers aged 2-5y and 6-14y, with little exposure from carriers aged younger than two years of age. We found that pneumococcal carriage was associated with the number of household members aged <5y, a recent cough, the total number of physical contacts in all age groups, and with stunting in children aged <5y. We estimate that all PCVs cover a substantial proportion of serotypes likely causing IPD in this population.

While we did not find local evidence of a significant association for all, many risk factors previously found to be associated with pneumococcal carriage are present in this population (25). Residents in Digaale live in overcrowded conditions and likely experience high levels of indoor air pollution. On average, one in five children are malnourished, and residents report a high frequency of direct contacts involving physical touch (12,26,27). While carriage prevalence was high, it is similar to that observed in non-displaced populations in other high-transmission settings in east Africa, and not as high as prevalence observed in rural Gambia where high carriage prevalence is sustained into older adulthood (28–31). Despite a high disease burden, displaced populations are understudied, and we are only aware of one other published carriage survey conducted in Mae La, a long-term camp for displaced people in Thailand, where carriage prevalence was estimated at a similar 80% in children <2y (7).

The most prevalent serotypes in Digaale (6B, 19F, and 23F) have often been observed to dominate carriage in other PCV-naïve populations, although the relatively high prevalence of 6C and low prevalence of 6A and 19A in our study is unusual (28–30,32). Around 50% of serotypes we detected were VTs, and the prevalence of observed serotypes included in the 10-valent Synflorix and PNEUMOSIL PCVs were similar. The proportion of VTs increased with valency of the vaccine. However, due to our relatively small sample size, serotype-specific confidence intervals are very wide. We estimate that any of the five PCVs are likely to cover the serotypes causing the majority of the pneumococcal disease burden. While serotype replacement would mitigate the overall PCV impact, substantial reductions in pneumococcal disease have been observed where PCVs have been introduced with sufficient coverage (33,34). While we did not collect data on the pneumococcal disease burden in Digaale, 43% of children under two years of age were reported to have been diagnosed with pneumonia in the six months preceding the survey (12), and pneumococci were one of the leading causes of childhood pneumonia globally in the pre-PCV era (35).

The combined contact and prevalence estimates showed that pneumococcal transmission in the <2y in Digaale was mostly driven by children aged 2-5y (39%) and 6-14y (25%), with little contribution to transmission from children younger than 2y old who have fewer social contacts. This could be important when designing vaccine strategies, especially those that partially rely on controlling pneumococcal transmission by indirect effects or need to prolong campaign effects in settings where continued vaccination through routine EPI schedule is not possible, as this requires extending the age range of the eligible population (2). While Digaale is an established camp that is safe and easy to access, this is not the case for many other displaced populations. In conflict settings, it is often not feasible to introduce routine immunization, and policy makers may choose alternative strategies that aim to immunize the subgroups that drive transmission, thereby indirectly protecting other subgroups at highest risk of severe disease.

Participants reported high rates of antibiotic usage in the two weeks preceding sample collection. This may be associated with the high proportion of participants with respiratory symptoms in the same period. However, we cannot rule out reporting bias. We found no association of antibiotic use with reduced carriage contrary to findings in other settings (29,36,37). Although in this study we do not have estimates of phenotypic pneumococcal resistance, microarray testing identified genes typically associated with pneumococcal resistance in a third of all samples, which may be consistent with high antibiotic pressure. The *tetM* gene, known to encode tetracycline resistance, was identified in 28% of pneumococci, mirroring its high prevalence in other studies (38,39). The *ermB* gene was found in 9% of pneumococci, and is associated with macrolide resistance (40). Future improved understanding of antimicrobial resistance in pneumococci would be useful to better understand the impact of more clinically-relevant antibiotics for standard care as well as the potential impact of mass-drug administration campaigns, a potential alternative intervention to reduce the pneumococcal disease burden proposed for crisis settings (41).

We assessed the relationship between a number of known risk factors with the odds of pneumococcal carriage and the mean pneumococcal carriage density. We found relationships in the expected direction for some risk-factors, such as an increased odds of carriage for participants with a higher number of household members under 5y of age, those with more direct contacts, and those with recent respiratory symptoms. Asymptomatic pneumococcal carriage has previously been found to be associated with rhinitis, and may be affected by other respiratory infections (42). It should be noted that confidence intervals were very wide due to a relatively low sample size and low variability within many risk factors. Moreover, we only adjusted our estimates for age and gender, and they are likely affected by residual confounding, while the large number of significance tests means that some spurious associations may have been estimated.

There are several limitations to our study. The study population was substantially smaller than expected as many shelters were uninhabited at the time of the study. Thus, we only reached 65% of our target sample size of 700 participants, particularly in young children, where we only reached 24% and 43% of our target sample size of 100 each in children aged <1 and 1y (12). We therefore pooled the <1 and 1y age groups in a single <2y age group, which allowed us to estimate age-specific prevalence with sufficient precision, but a larger sample would have resulted in more detailed estimates. We could only conduct data collection during daylight hours and may have missed older individuals who work outside Digaale, as many leave the camp very early in the morning and return late at night. This is likely reflected in the gender distribution of the recruited sample, but unlikely to have affected our carriage estimates, as prevalence is low in these older age groups and unlikely to differ substantially from those who were present in Digaale. We post-stratified our estimates to adjust for any imbalances in our sample and did not detect differences. Although, pneumococcal carriage is generally consistent across seasons (43), we only conducted a single cross-sectional survey and do not know how estimates may change throughout the year. Carriage prevalence was similar to that in general populations in East Africa, but no other prevalence estimates exist for Somaliland, and we cannot infer how results may differ from the general Somaliland population. Our study was not powered to detect relationships between carriage and risk factors, which may explain why we did not find statistically significant effects in most univariate analyses, and a prospective cohort design would be more suitable to infer causality. Finally, many risk factors were self-reported, and their accuracy may be affected by reporting bias.

Ideally, pneumococci are stored at ULT to maintain long-time viability, but we experienced several challenges related to sample storage and shipment. Sample shipment was substantially delayed, partly due to the COVID-19 pandemic, and after several months of storage at ULT, swabs had to be temporarily transferred to a −20°C freezer to allow for ULT freezer repairs. We were not able to transport samples at ULT as local airlines did not accept shipments of dry ice. However, we have shown separately that effect on pneumococcal viability is limited if stored at −20°C for up to three weeks (15), which was maintained in our study in the periods that ULT storage was not feasible. We further monitored sample viability by incorporation of *lytA* qPCR, a molecular screening assay that is not expected to be affected by culture viability, and did not observe a large number of non-culturable samples that were *lytA* positive. Despite transit delays during the second shipment of most of our samples during which temperatures may have exceeded 20°C for up to 2.5 days, we found no difference between carriage and VT prevalence estimates between these samples and those transported during the first shipment. Hence, we believe any effect on sample viability was limited and did not greatly affected our results, supported by the ability to culture at high prevalence and with detection of serotypes carried at low abundance.

## Conclusion

We found high pneumococcal carriage prevalence in a PCV-naïve population living in an IDP camp in Somaliland, consistent with carriage rates in non-displaced populations in other high transmission settings. About half of all circulating pneumococci were included in currently available PCVs. We estimate that at least half of all resulting IPD cases in this population were caused by serotypes included in PCVs, indicating the potential for substantial vaccine effects. Transmission was primarily driven by children 2-5 years and 6-14 years old, partially exceeding the proposed age eligibility for PCV campaigns that aim to temporarily reduce transmission in crisis-affected populations. These findings advance our understanding of pneumococcal carriage in crisis-affected populations and provide important evidence for the design of future vaccination strategies.

## Data Availability

All data produced in the present study are available upon reasonable request to the authors

## Acknowledgements

We sincerely thank the Digaale community who participated in this study, as well as Hamse Shaban Ahmed, Ayaan Ismail Adan, Khadar Abiib Ahmed, Ayaan Mohamoud Ali, Hamda Awil Garaad, Ahmed Yusuf Mohamed, Suaad Abdi Osman, Amiin Abdi Ismail, Nimco Abdilahi Ismail, AbdiFatah Mohamed Ahmed, Mustafe Shugri Mohamed, and Nimco Mohamed Abdi from the Republic of Somaliland Ministry of Health Development for their data collection efforts. Electronic data solutions were provided by LSHTM Open Data Kit (odk.lshtm.ac.uk)

