## Supplemental Material for "Pre-vaccination carriage prevalence of *Streptococcus pneumoniae* serotypes among internally displaced people in Somaliland"

Additional analysis scripts and anonymized data will be made available on GitHub:

https://github.com/kevinvzandvoort/espicc_somaliland_digaale_carriage_survey_2019.

**TABLE OF CONTENTS**

Section A. Data collection and matching datasets ... 1
Section B. Sample shipment ... 4
*Passive temperature-controlled shipment* ... 4
*Temperature during sample shipments* ... 5
*Assessing carriage between shipments* ... 6
Section C. Additional microbiological analyses ... 9
*Multiple serotype carriage, density, and resistant genes* ... 9
*Association between risk factors and pneumococcal density* ... 10
*Impact of weighting on serotype distribution* ... 12
*Serotype distribution by age* ... 14
*Carriage prevalence by age* ... 15
*Assessing the choice of post-stratification weights on prevalence estimates* ... 16
*Carriage prevalence compared to other settings* ... 18
*Contribution of different age groups towards the age-specific exposure to pneumococcus* ... 19
Section D. Invasive pneumococcal disease estimates ... 20
*Age- and serotype-specific invasiveness* ... 20
*Estimating invasiveness in Digaale* ... 23
*Results* ... 24
References used in Supplemental Material ... 27

### Section A. Data collection and matching datasets

Data was collected in multiple stages and using multiple Open Data Kit (ODK) forms. All shelters in Digaale IDP camp were visited. In shelters where at least one adult was present who consented to participation, a household survey was conducted using a household survey form which collected data on the household level (stored in the *household* dataset) and individual demographic data for all household members living in the household (stored in the *household members* dataset). Quota sampling was used to invite individual household members to participate in the contact survey, and data collectors returned to the household to conduct the contact survey two days after the household survey. For participants in the contact survey aged 6m to 59m, a nutritional assessment was conducted immediately after the contact survey. These data were collected on separate contact survey and nutrition forms, and stored in the *contact* and *nutrition* datasets, respectively. Finally, data collectors returned to all contact survey participants in the final two weeks of the survey, and asked them to have a nasopharyngeal swab taken. Swab-related data were collected using a swab form, and subsequently stored in the *swab* dataset. Additional household members who were not initially selected for participation in the contact survey were sampled for participation in nasopharyngeal swab selection (and a nutritional assessment, for age-eligible children).

Unique household and individual identifiers were automatically assigned in the household form, and manually re-entered in subsequent forms. Due to human error, some identifiers were incorrectly re-entered in subsequent forms, resulting in unmatched data between datasets for some records.

Supplemental Figure A1 shows an UpSet plot (1) with the number of matched records between multiple datasets. The majority (400; 88%) of records for collected swabs could be matched between all datasets for which forms were completed: i) 243 records with household, contact, and swab data for people age-ineligible for the nutritional assessment, ii)

112 records with household, contact, nutrition, and swab data for people age-eligible for the nutritional assessment, and iii) 45 records with household and swab data for people age-ineligible for the nutritional assessment who were not invited for the contact survey.

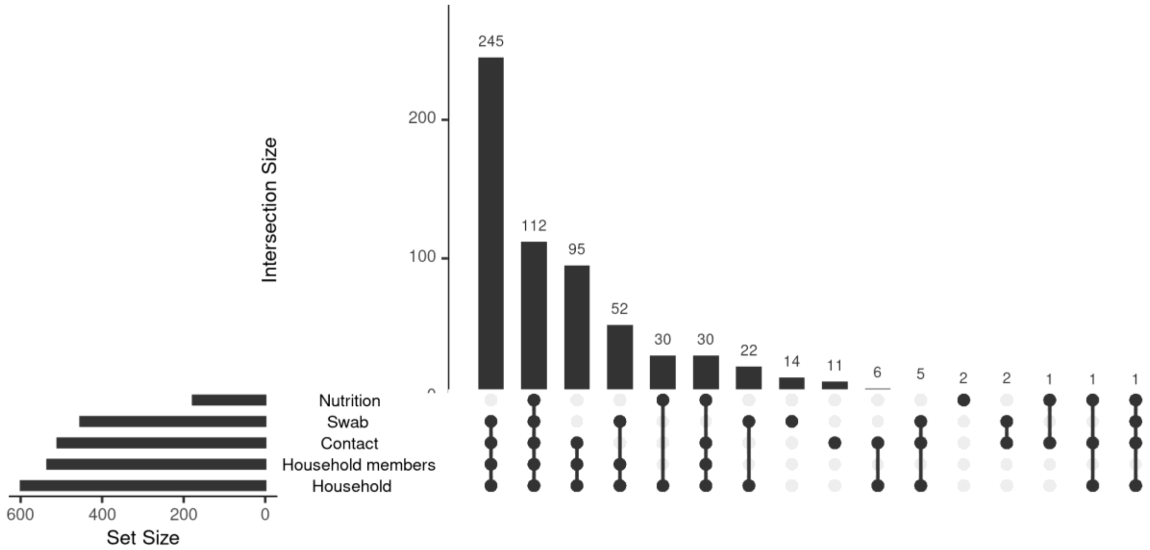

**SUPPLEMENTAL FIGURE A1. MATCHING RECORDS BETWEEN DATASETS.**
UpSet plot showing the number of matching records between the different datasets, matches show the number of records that can be linked to the same person in the household member, contact, nutrition, and swab datasets, and to the same household in the household dataset. Data collected for households and household members that did not participate in an individual survey is not shown. Nutrition data is only collected for participants aged 6 to 59 months.

There were 53 records for collected swabs that could not be completely matched between all datasets for which forms were completed: i) there were two records for people age-eligible for the nutritional assessment, for who records could be linked to the household, contact, and swab, dataset, but not the nutrition dataset; ii) there were seven records for collected swabs that could be linked to household and swab data for people age-eligible for the nutritional assessment, that could not be linked to records in the nutrition dataset; iii) there were 22 records for collected swabs that could be linked to records for an individual household, but not to records for an individual in any of the other datasets, including household members; iv) 14 records for swabs could not be linked to any of the other datasets; v) 5 could be linked to an individual record in the contact dataset and to records for an individual household, but not to records for individual household members, and two of these were age-eligible for the nutritional assessment; vi) there were two records that could only be linked to records in the contact dataset; and vii) there was one record that could be linked to all but the household members datasets.

The inability to match certain records prevented the inclusion of data from some swabs in certain analyses.  resulting in missing data for some covariates. Future surveys should aim to minimize challenges with matching identifiers.

#### Section B. Sample shipment

##### *Passive temperature-controlled shipment*

Due to logistical challenges, we were not able to ship collected samples on ultra-low-temperatures (ULT), which is the gold standard. Instead, samples were shipped using Thermocon Classic 15, a prequalified shipping solution utilizing phase change materials (PCMs), that provided passive cooling to keep temperatures at below -15°C for up to 96h as per manufacturer instructions (Schaumaplast, DE).

Manufacturer instructions state to precondition PCMs at -20°C prior to shipment. Instead, we tested preconditioning of PCMs at -70°C in a ULT freezer and measured the temperature of the shipping solution during an empty test-shipment of the shipping solution from London to Hargeisa in November 2020 using a Testo 184 T4 (Testo, DE) temperature logger (Supplemental Figure B1 and Supplemental Table B1). Temperatures were maintained at below -15°C for up to 160h during this test shipment. Temperatures rose above 0°C after 204h. We did not measure the ambient temperatures during this shipment.

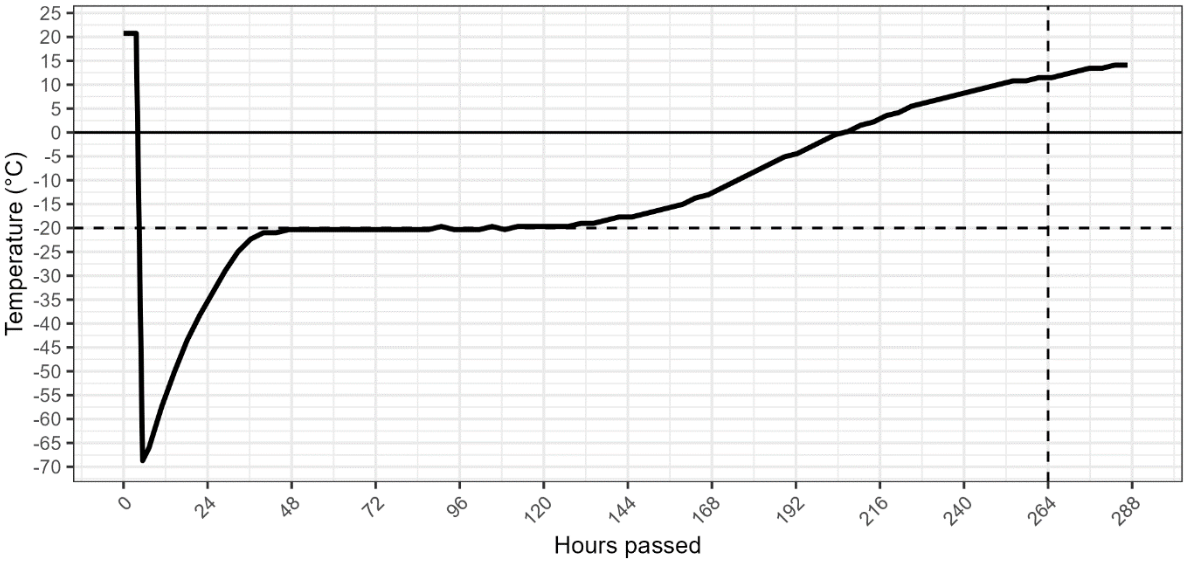

**SUPPLEMENTAL FIGURE B1. TEMPERATURE OF TEST SHIPMENT OVER TIME.**
Temperature (in degrees Celsius) inside a Thermocon Classic 15 over time, during a shipment from London to Hargeisa. The dotted vertical line shows the temperature after 264 hours.

**SUPPLEMENTAL TABLE B1. TIME UNTIL TEMPERATURE EXCEEDANCE.**

| Temperature exceeded | Hours passed | Days passed |
|---|---|---|
| -20 °C | 87 | 3.6 |
| -15 °C | 160 | 6.7 |
| -10 °C | 175 | 7.3 |
| -5 °C | 189 | 7.9 |
| 0 °C | 204 | 8.5 |
| 5 °C | 222 | 9.3 |
| 10 °C | 248 | 10.3 |
| 15 °C | 299 | 12.5 |

##### *Temperature during sample shipments*

We initially tested our shipment route using the PCMs with a pilot shipment of 81 samples in May 2021. Samples arrived in Nairobi, Kenya within 4 days, where they were placed in a -20°C freezer and shipped on dry ice after a further 3 days to the Murdoch Children's Research Institute (MCRI) in Melbourne, Australia for storage and analysis. Samples remained below -20°C for the entire duration of the pilot shipment (Supplemental Figure B2).

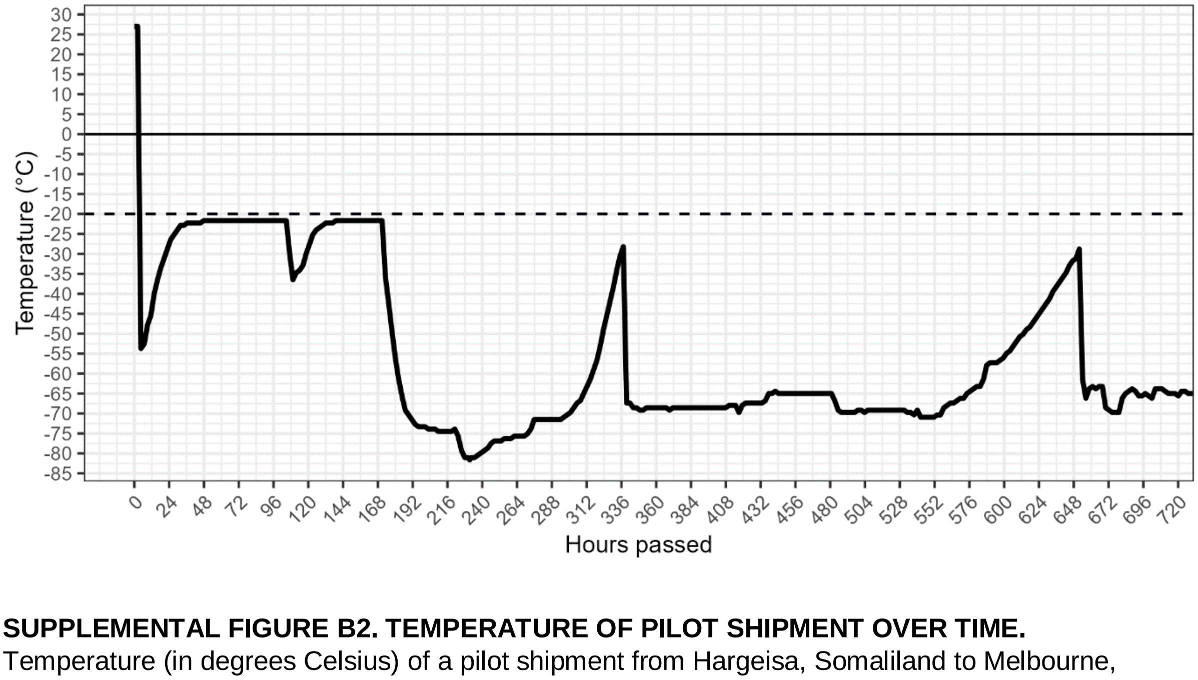

**SUPPLEMENTAL FIGURE B2. TEMPERATURE OF PILOT SHIPMENT OVER TIME.** Temperature (in degrees Celsius) of a pilot shipment from Hargeisa, Somaliland to Melbourne, Australia transiting through Nairobi, Kenya.

We subsequently shipped all remaining samples in December 2021. This shipment got delayed in transit to Nairobi, and samples were only temperature controlled through the passive cooling from PCMs for a total of 264h (11 days). No temperature data was available for this shipment. However, if temperatures during this shipment were similar as during the test shipment, this may indicate that the temperature of the swabs may have been >0°C for up to 2.5 days and may have risen to 12.5°C (Supplemental Figure B1). This extended transit, and potential rise in temperature, may have affected the viability of the samples. Pell et al. previously found that pneumococcal isolates at low density remained detectable when stored on flocked swabs in skim milk-tryptone-glucose glycerol medium (STGG) at -20°C for up to three weeks, though at reduced levels compared to the gold standard of ULT storage (2). They also found that isolates remained viable when stored in STGG at 4°C for up to 2 days, but that viability rapidly declined after >2 days.

*Assessing carriage between shipments*

To understand whether the extended period at which the 372 swabs transported during the second shipment were likely exposed to higher temperatures may have affected the viability of these samples, we compared the pneumococcal carriage rates estimated from these swabs to those of the 81 samples shipped during the first pilot shipment. Sample-specific poststratification weights on age and gender were used to calculate population level prevalence estimates.

Overall pneumococcal carriage prevalence was similar in estimates from the two shipments (Supplemental Table B2), at 39.6% (95%CI 28.2 – 51.0) compared to 35.2% (30.1 – 40.4) in all ages, and 69.9% (57.3 – 82.5) compared to 70.4% (63.0 – 77.8) in children <5y. There are some difference in the proportion of VT-covered serotype carriers, at 58.3% (38.7 – 77.8) compared to 42.2% (33.4 – 51.0) for PNEUMOSIL-covered serotype carriers in all age-groups, and 30.5% (5.6 – 55.5) compared to 60.7% (51.4 – 70.1) in children <5y. Due to the small sample sizes, uncertainties around those estimates are wide, and sampling error cannot be ruled out to explain those differences.

We assessed whether there was any statistical evidence of a difference in pneumococcal carriage between the samples shipped in the two samples. There was no evidence of a difference in the odds of pneumococcal carriage for samples shipped during the pilot shipment compared to those shipped in the second shipment (OR 1.14; 95%CI 0.69 – 1.85; p=0.61), or of a difference in the odds of carrying PNEUMOSIL VTs in those who carried pneumococci (OR: 0.70; 0.09 – 14.44; p=0.76), as assessed using a logistic regression model. There also was no evidence of a difference in the mean log10 density in samples with pneumococci (-0.25; -0.64 – 0.15; p=0.22), as assessed using a linear regression model. Finally, there also was no evidence of a relative difference in the total number of serotypes identified (0.86; 0.61 – 1.19; p=0.371), in those carrying at least one serotype, as assessed using a Poisson regression model.

**SUPPLEMENTAL TABLE B2. CARRIAGE PREVALENCE IN PILOT AND SECOND SHIPMENT**

| **Variable** | **Obs**[a] | **Sample value** | **Population est**[b] | | **Population est (<5y)**[b] | |
|---|---|---|---|---|---|---|
| *Pilot shipment* | | | | | | |
| Overall carriage | 34/81 | 42.0% | 39.6% | 28.2 - 51.0 | 69.9% | 57.3 - 82.5 |
| PNEUMOSIL-covered serotype carriers | 15/34 | 44.1% | 58.3% | 38.7 - 77.8 | 30.5% | 5.6 - 55.5 |
| Synflorix-covered serotype carriers | 16/34 | 47.1% | 56.0% | 36.1 - 75.9 | 32.7% | 7.1 - 58.3 |
| Prevenar 13-covered serotype carriers | 19/34 | 55.9% | 46.8% | 26.2 - 67.4 | 27.3% | 2.6 - 52.1 |
| Vaxneuvance-covered serotype carriers | 19/34 | 55.9% | 46.8% | 26.2 - 67.4 | 27.3% | 2.6 - 52.1 |
| Prevenar 20-covered serotype carriers | 22/34 | 64.7% | 41.1% | 20.5 - 61.7 | 20.1% | 0.0 - 43.5 |
| *Second shipment* | | | | | | |
| Pneumococcal carriage | 144/370 | 38.9% | 35.2% | 30.1 - 40.4 | 70.4% | 63.0 - 77.8 |
| PNEUMOSIL-covered serotype carriers | 72/144 | 50.0% | 42.2% | 33.4 - 51.0 | 60.7% | 51.4 - 70.1 |
| Synflorix-covered serotype carriers | 70/144 | 48.6% | 40.4% | 31.7 - 49.2 | 58.6% | 49.0 - 68.1 |
| Prevenar 13-covered serotype carriers | 81/144 | 56.2% | 50.8% | 42.0 - 59.6 | 62.2% | 53.0 - 71.5 |
| Vaxneuvance-covered serotype carriers | 81/144 | 56.2% | 50.8% | 42.0 - 59.6 | 62.2% | 53.0 - 71.5 |

**SUPPLEMENTAL TABLE B2. CARRIAGE PREVALENCE IN PILOT AND SECOND SHIPMENT**

| Variable | Obs[a] | Sample value | Population est[b] | | Population est (<5y)[b] | |
|---|---|---|---|---|---|---|
| Prevenar 20-covered serotype carriers | 90/144 | 62.5% | 57.9% | 49.1 - 66.8 | 66.4% | 57.2 - 75.6 |

### Section C. Additional microbiological analyses

#### *Multiple serotype carriage, density, and resistant genes*

The median number of serotypes in samples in which pneumococci were detected by microarray was 1 (IQR 1 – 2). In 30% (95%CI 23 – 37) of these samples, more than one serotype was detected (Supplemental Table C1). The median density was 6.5 (IQR 5.4 – 7.0) $log_{10}$ GE/ml for all samples. In 30% (21 – 41) of pneumococci, at least one resistance gene was detected. The most common resistant gene was *tetM*, present in 28% (19 – 39) of pneumococci. Samples in which more than one pneumococcal serotype, or in which other species were detected, were excluded from estimates of the proportion with resistant genes.

**SUPPLEMENTAL TABLE C1. OTHER MICROBIOLOGICAL RESULTS**

| Variable | Obs | Sample value | |
|---|---|---|---|
| *Multiple serotype carriage* | | | |
| Median number of serotypes[b] | 176 | 1 | 1 - 2 (IQR) |
| Multiple serotype carriage | 52/176 | 30% | 23 - 37 |
| *Pneumococcal density (log10 GE/ml; median)*[a] | | | |
| All samples | 176 | 6.45 | 5.38 - 7.00 (IQR) |
| Samples with only 1 serotype | 124 | 6.43 | 5.32 - 6.93 (IQR) |
| Samples with > 1 serotype | 52 | 6.58 | 5.67 - 7.06 (IQR) |
| Per serotype | 248 | 6.07 | 5.02 - 6.84 (IQR) |
| *Other species detected by microarray* | | | |
| All | 63/191 | 33% | . |
| *Resistant genes*[c] | | | |
| Any | 28/92 | 30% | 21 - 41 |
| Gene-specific | | | |
| *aphA3* | 1/92 | 1% | 0 - 6 |
| *cat* | 1/92 | 1% | 0 - 6 |
| *ermB* | 8/92 | 9% | 4 - 16 |
| *ermC* | 0/92 | 0% | 0 - 4 |
| *mefA* | 2/92 | 2% | 0 - 8 |
| *tetK* | 1/92 | 1% | 0 - 6 |
| *tetL* | 0/92 | 0% | 0 - 4 |
| *tetM* | 26/92 | 28% | 19 - 39 |
| *tetO* | 0/92 | 0% | 0 - 4 |
| *sat4* | 1/92 | 1% | 0 - 6 |

a. Median and IQR of log10 transformed density estimates.
b. In those who carry pneumococci
c. Restricted to samples with only one pneumococci, and no other species detected

The odds that an individual serotype was the dominant serotype among all carried serotypes in a sample was 2.0 (1.1 – 3.7) times higher for VTs than for NVTs. The odds were similar when restricting the dataset to samples in which both VTs and NVTs were detected, these odds increased to 2.8 (1.0 – 8.0).

**SUPPLEMENTAL TABLE C2. ASSOCIATION BETWEEN SEROTYPE AND DOMINANT CARRIAGE**

| Variable | OR | 95% CI | p-value | N |
|---|---|---|---|---|
| *Serotype is dominant (all participants)* | | | | |
| NVT | 1.00 | | | 248 |
| VT | 2.00 | 1.10 - 3.70 | 0.020 | 248 |
| *Serotype is dominant (in those carrying both VT and NVT)* | | | | |
| NVT | 1.00 | | | 65 |
| VT | 2.80 | 1.00 - 8.00 | 0.051 | 65 |

##### *Association between risk factors and pneumococcal density*

We used linear regression to assess the association between observed risk factors and the mean log10 density of pneumococcal serotypes (Supplemental Table C3). We found very weak evidence that living with one additional household member aged <5y was associated with a 0.18 (95%CI -0.01 – 0.37) increase in mean log10 density, and for an association between having a sore throat in the 2 weeks preceding the survey and a 0.40 reduction (0.00 – 0.79) in mean log10 density. There was some very weak evidence for a reduction in mean log10 density for pneumococci carried by children with improved weight-for-height z-scores, but no evidence for an association with any other potential risk factor.

**SUPPLEMENTAL TABLE C3. ASSOCIATION BETWEEN RISK FACTORS AND PNEUMOCOCCAL DENSITY**

| Variable | Mean difference[a,b] | 95% CI | p-value | N[c] |
|---|---|---|---|---|
| *Demographic characteristics* | | | | |
| Household size | -0.02 | -0.10 - 0.05 | 0.533 | 172 |
| Household members <5y | 0.18 | -0.01 - 0.37 | 0.064 | 172 |
| *Shelter quality* | | | | |
| House leakage | | | | |
| no | *ref* | | | |
| yes | -0.19 | -0.56 - 0.18 | 0.307 | 172 |
| House draft | | | | |
| no | *ref* | | | |

**SUPPLEMENTAL TABLE C3. ASSOCIATION BETWEEN RISK FACTORS AND PNEUMOCOCCAL DENSITY**

| Variable | Mean difference[a,b] | 95% CI | p-value | N[c] |
|---|---|---|---|---|
| yes | 0.27 | -0.07 - 0.61 | 0.116 | 172 |
| *Indoor air pollution* | | | | |
| Fuel firewood | | | | |
| no | *ref* | | | |
| yes | 0.02 | -0.41 - 0.45 | 0.934 | 172 |
| Fuel charcoal | | | | |
| no | *ref* | | | |
| yes | -0.06 | -0.39 - 0.27 | 0.713 | 172 |
| Ventilation | | | | |
| no | *ref* | | | |
| yes | 0.23 | -0.32 - 0.78 | | 172 |
| cook outside | 0.08 | -0.41 - 0.57 | 0.628 | 172 |
| *Current health[d]* | | | | |
| Antibiotic use | | | | |
| no | *ref* | | | |
| yes | 0.09 | -0.23 - 0.42 | 0.585 | 163 |
| Respiratory symptoms | | | | |
| no | *ref* | | | |
| yes | -0.10 | -0.46 - 0.26 | 0.587 | 164 |
| Cough | | | | |
| no | *ref* | | | |
| yes | 0.05 | -0.27 - 0.36 | 0.778 | 164 |
| Sore throat | | | | |
| no | *ref* | | | |
| yes | -0.40 | -0.79 - 0.00 | 0.050 | 164 |
| Headache | | | | |
| no | *ref* | | | |
| yes | -0.17 | -0.63 - 0.28 | 0.460 | 164 |
| Fever | | | | |
| no | *ref* | | | |
| yes | -0.17 | -0.50 - 0.17 | 0.340 | 164 |
| Diarrhoea | | | | |
| no | *ref* | | | |
| yes | -0.06 | -0.50 - 0.38 | 0.793 | 164 |
| *Morbidities[e]* | | | | |
| Pneumonia 6m[f] | | | | |
| no | *ref* | | | |
| yes | 0.12 | -0.26 - 0.51 | 0.528 | 152 |
| Sickle Cell | | | | |
| no | *ref* | | | |
| yes | 0.10 | -0.76 - 0.96 | 0.826 | 152 |
| Asthma | | | | |
| no | *ref* | | | |
| yes | 1.49 | -0.02 - 3.01 | 0.056 | 152 |
| Diabetes | | | | |
| no | *ref* | | | |

**SUPPLEMENTAL TABLE C3. ASSOCIATION BETWEEN RISK FACTORS AND PNEUMOCOCCAL DENSITY**

| Variable | Mean difference[a,b] | 95% CI | p-value | N[c] |
|---|---|---|---|---|
| yes | 1.53 | -0.61 - 3.67 | 0.163 | 152 |
| *Individual substance use* | | | | |
| Tobacco | | | | |
| no | *ref* | | | |
| yes | -0.20 | -2.37 - 1.98 | 0.861 | 152 |
| Khat | | | | |
| no | *ref* | | | |
| yes | -0.20 | -2.37 - 1.98 | 0.861 | 152 |
| *Household substance use[g]* | | | | |
| Household smoke | | | | |
| no | *ref* | | | |
| yes | -0.09 | -0.43 - 0.25 | 0.602 | 172 |
| Household snuff | | | | |
| no | *ref* | | | |
| yes | 0.02 | -0.54 - 0.58 | 0.949 | 172 |
| Household khat | | | | |
| no | *ref* | | | |
| yes | -0.26 | -0.58 - 0.06 | 0.107 | 172 |
| *Contact behaviour* | | | | |
| Total number of direct contacts | -0.03 | -0.08 - 0.02 | 0.303 | 151 |
| Total number of physical contacts | -0.01 | -0.06 - 0.04 | 0.602 | 151 |
| *Malnutrition in <5y* | | | | |
| Weight-for-age z-score | 0.10 | -0.12 - 0.33 | 0.358 | 81 |
| Weight-for-height z-score | 0.16 | -0.02 - 0.35 | 0.088 | 81 |
| Height-for-age z-score | -0.05 | -0.23 - 0.12 | 0.556 | 81 |
| MUAC[h]-for-age z-score | 0.01 | -0.23 - 0.25 | 0.933 | 81 |
| MUAC[h] (in cm) | 0.02 | -0.18 - 0.23 | 0.823 | 81 |

a. Density estimates were log10 transformed prior to linear regression.
b. Estimates are adjusted for age and gender.
c. Total number of records used in regression.
d. Self-reported antibiotic use and symptoms in 2 weeks preceding the survey.
e. Self-reported diagnosed morbidities.
f. Pneumonia diagnosis in the 6m preceding the survey.
g. Substance use by at least one household member.
h. Middle Upper Arm Circumference

#### *Impact of weighting on serotype distribution*

In our main analysis, we weighted the pneumococcal serotype distribution by their relative abundance. We assessed the sensitivity to this approach by comparing it to the serotype distribution of only dominant serotypes, and all serotypes (unweighted). There were no major differences between the three distributions (Supplemental Figure C1), with serotypes 6B, 19F, and 23F as the most common serotypes in all three.

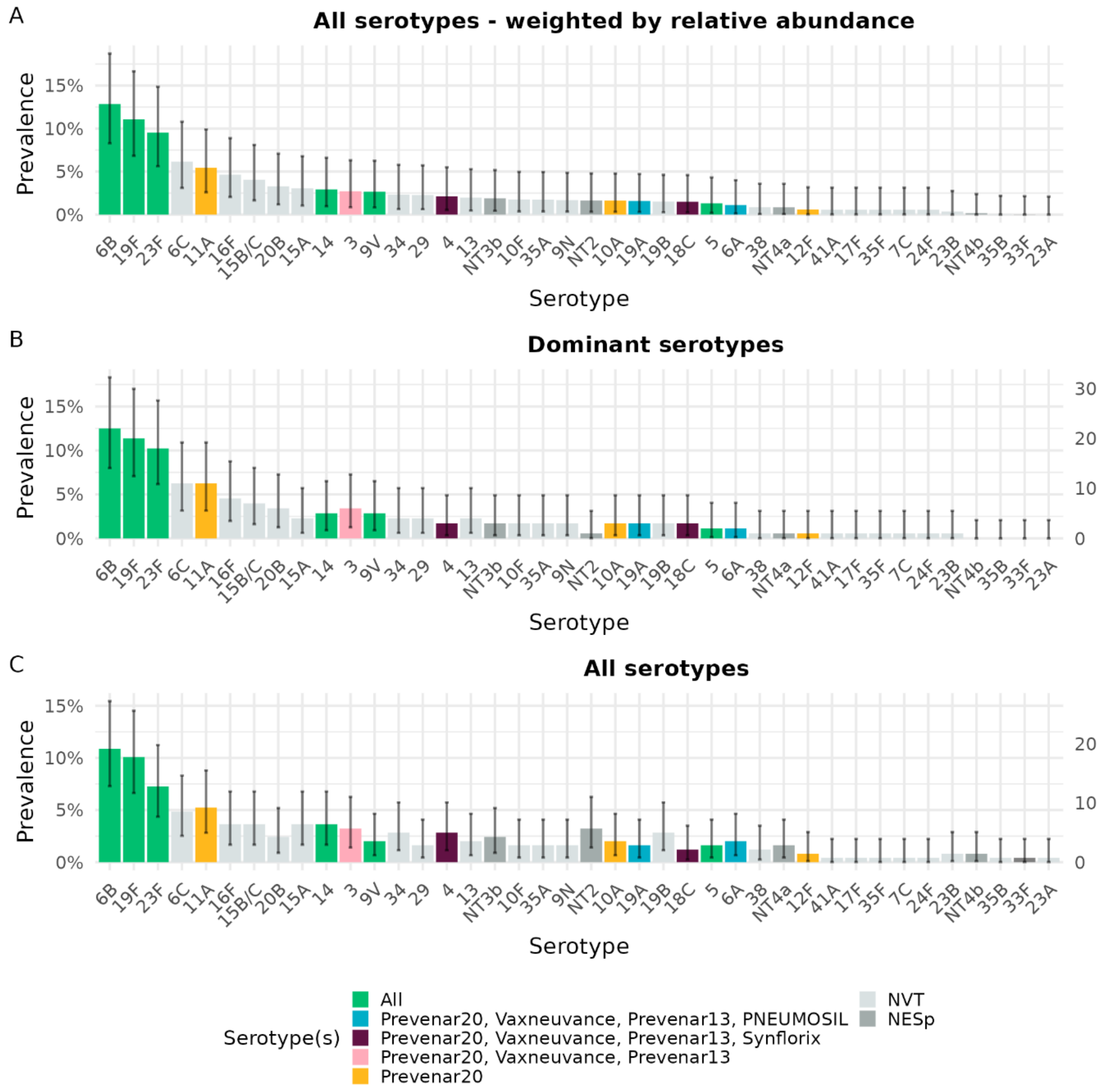

**SUPPLEMENTAL FIGURE C1. PNEUMOCOCCAL SEROTYPE DISTRIBUTION.**
Bars show the proportion of serotypes among all identified pneumococci in Digaale IDP camp. A: serotype distribution of all serotypes, weighted by their relative abundance of carriage; B: serotype distribution of dominant serotypes only; C: unweighted serotype distribution of all serotypes. Coloured bars show the serotypes included in the three licensed PCVs, dark grey bars non-encapsulated pneumococci, and light grey bars other serotypes not included in the PCVs. Error bars show 95% confidence intervals for each estimate.

*Serotype distribution by age*

We also stratified the serotype distribution by age, in those carried by children <5y, and by people aged ≥5y (Supplemental Figure C2). The serotype distribution differed substantially between age groups, although confidence intervals around the point estimates are wide. In children <5y, the five most common serotypes were 6B (10%; 95%CI 6 – 16), 19F (10%; 6 – 16), 23F (7%; 4 – 12), 11A (4%; 2 – 8), and 6C (3%; 1 – 7). In contrast, the relative proportion of these serotypes substantially decreased in people ≥5y, with a much more uniform distribution. In these older people, the five most common serotypes were 15B/C (3%; 1 – 7), 6B (3% 1 – 7), 6C (3%, 1 – 7), 3 (2%, 1 – 6), and 16F (2% 1 – 6).

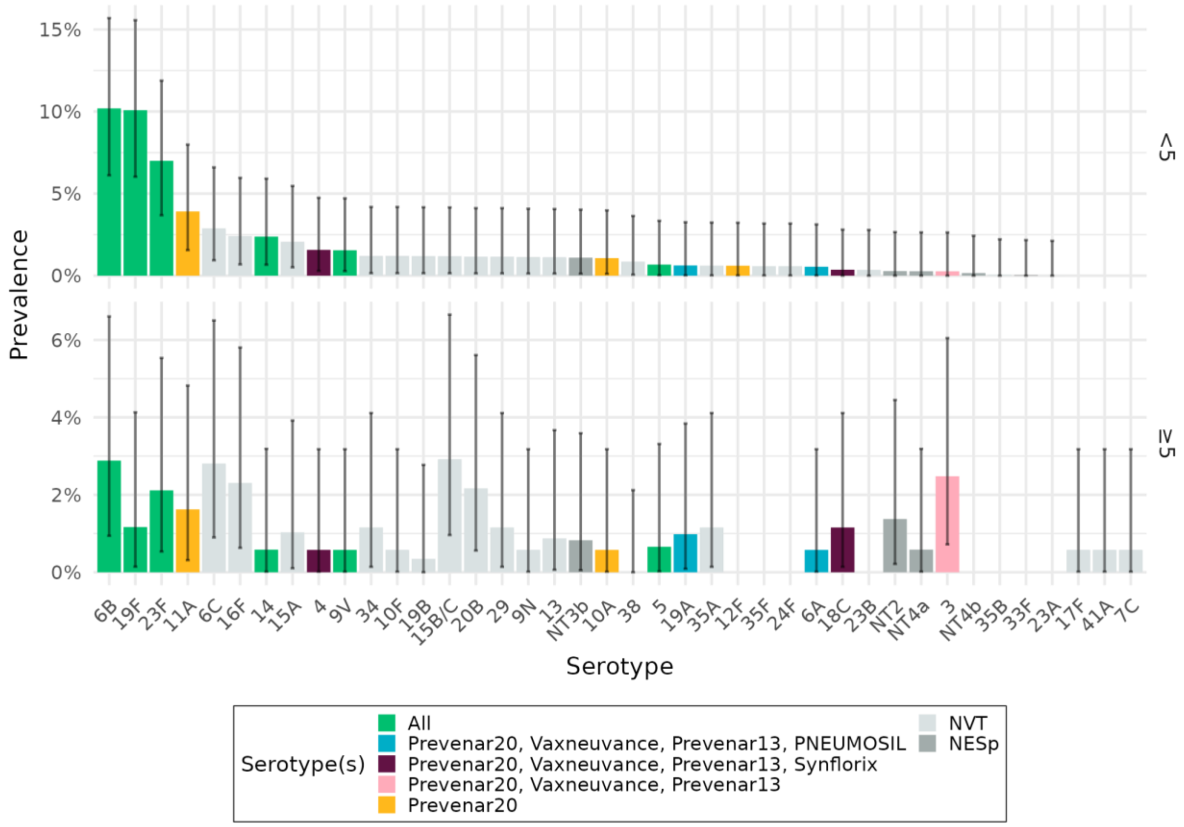

**SUPPLEMENTAL FIGURE C2. PNEUMOCOCCAL SEROTYPE DISTRIBUTION.**
Bars show the proportion of serotypes among all identified pneumococci in Digaale IDP camp, weighted by their relative abundance, in participants aged <5y (A), and ≥5y (B). Coloured bars show the serotypes included in the three licensed PCVs, dark grey bars non-encapsulated pneumococci, and light grey bars other serotypes not included in the PCVs. Error bars show 95% confidence intervals for each estimate.

*Carriage prevalence by age*

Supplemental Figure C3 shows overall pneumococcal prevalence, and prevalence of VTs, NVTs, and NESp by age, for different PCV formulations. We also show the distributions for dominant serotype carriage only.

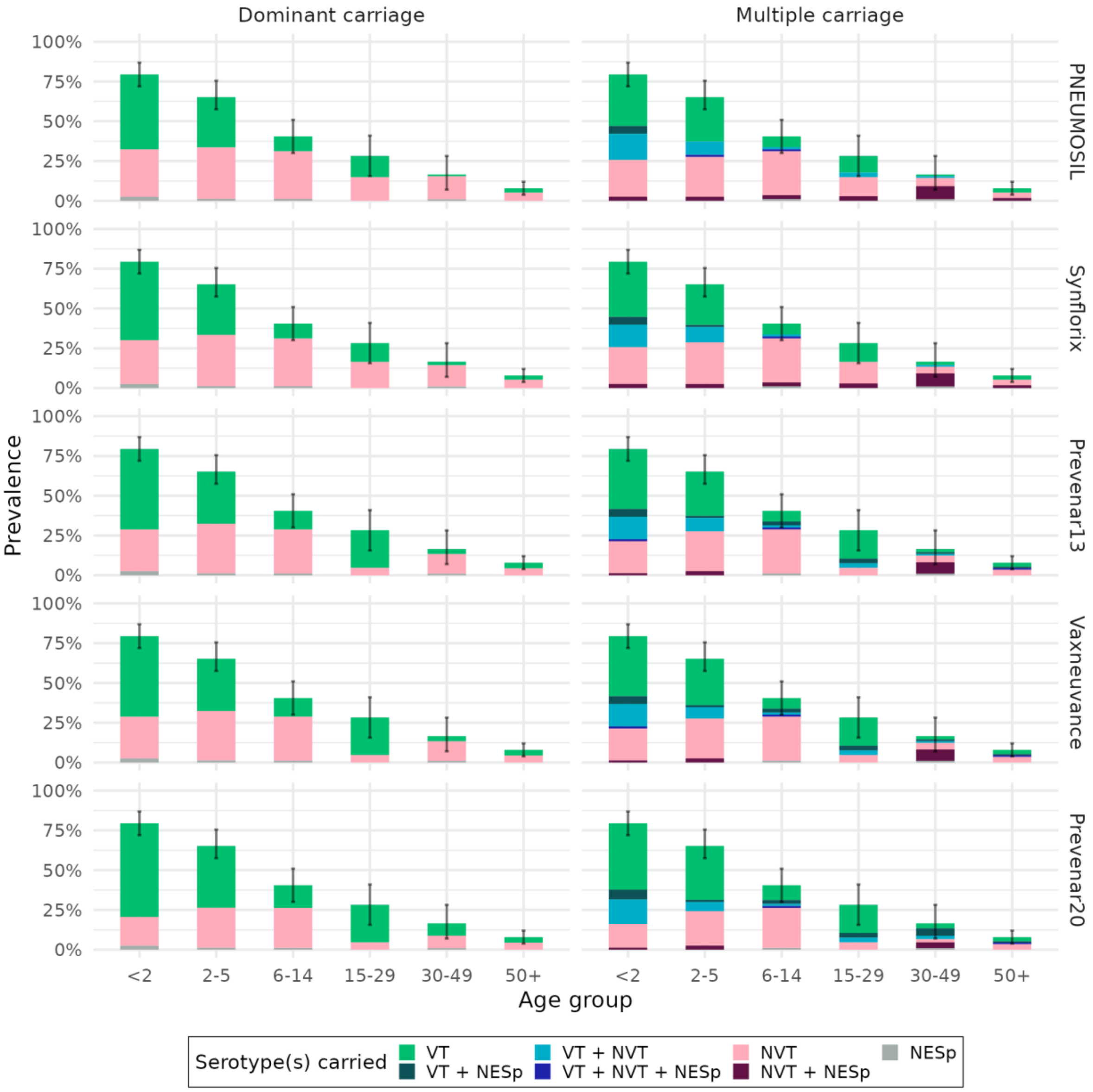

**SUPPLEMENTAL FIGURE C3. PREVALENCE AND SEROTYPE DISTRIBUTION BY AGE.** Facet columns show the serotype distribution of dominant serotypes only, and of all serotypes in multiple serotype carriers. Facet rows use different definitions for vaccine types: PNEUMOSIL, Synflorix, Prevenar 13, Vaxneuvance, Prevenar 20. Bars show the estimated prevalence of pneumococcal serotypes by age group, weighted for age and gender. Error bars show 95% confidence intervals around overall pneumococcal carriage prevalence. Colours show the prevalence of serotypes that are carried; VT: only vaccine type(s), NVT: only non-vaccine type(s); NT: only non-encapsulated type(s); and where applicable VT + NVT: both vaccine- and non-vaccine type(s). Multiple carriage with non-encapsulated type(s) is shown as a darker shading.

*Assessing the choice of post-stratification weights on prevalence estimates*

As participants were not selected using a stratified random sampling design, we post-stratified our results to calculate population-level estimates. Data were stratified by gender (male or female) and the following age groups: <2, 2-5, 6-14, 15-29, 30-49, 50+ years of age, and poststratification was implemented using the *Survey* package in R (3). Prevalence estimates in our main manuscript are weighted by the calculated poststratification weights on age and gender, unless otherwise stated.

We assessed the sensitivity to the choice of variables used to weight the data by comparing prevalence estimates weighted by the variables listed in Supplemental Table C4.

**SUPPLEMENTAL TABLE C4.** Post-stratification weights used to calculate population-level estimates.

| Weights | Age group [a] | Gender [b] | Household size [c] |
|---|---|---|---|
| I [d] | ✓ | ✓ | × |
| II | ✓ | × | ✓ |
| III | ✓ | × | × |
| IV | × | × | × |

*Variables used in calculating the weights used to construct contact intensities*

a. *Categorised as <2, 2-5, 6-14, 15-29, 30-49, 50+ years of age*
b. *Female or male*
c. *Categorised by quantiles: 1-2, 3-4, 5-6, and 7-12 household members*
d. *Main weights used in the analysis*

There was little difference in the estimated pneumococcal prevalence by different post-stratification weights. The unweighted sample estimate of 39.5% (35.3 – 43.6) was adjusted to prevalence ranging between 35.0% – 35.8%, while the unweighted sample estimate of 71.7% in children <5y did not substantially change, with weighted estimates ranging between 70.0% – 70.4%.

**SUPPLEMENTAL TABLE C5. Pneumococcal prevalence estimates by different post-stratification weights.**

| Weights | Obs[a] | Sample value | Population est | | Population est (<5y) | |
|---|---|---|---|---|---|---|
| I | 175/445 | 39.3% | 35.8% | 31.1 - 40.4 | 70.0% | 63.7 - 76.4 |
| II | 174/443 | 39.3% | 35.4% | 30.8 - 40.1 | 70.4% | 64.0 - 76.8 |
| III | 175/445 | 39.3% | 35.0% | 30.6 - 39.4 | 70.0% | 63.5 - 76.5 |
| IV | 178/451 | 39.5% | 39.5%[b] | 35.3 - 43.6 | 71.7% [b] | 65.8 - 77.6 |

a. Number of observations included in sample. Observations with missing values for variables used in weighting are excluded from the dataset.

| SUPPLEMENTAL TABLE C5. Pneumococcal prevalence estimates by different post-stratification weights. | | | | |
|---|---|---|---|---|
| **Weights** | **Obs[a]** | **Sample value** | **Population est** | **Population est (<5y)** |

b. Population estimate is the same as the sample estimate, as estimates are unweighted.

There were also no substantial differences in the serotype distribution by age using different weights. The most prominent difference was an increase in the proportion of NVTs and NTs co-carried in people aged 30-49 when weighting on age and gender, but overall prevalence in this age groups is small.

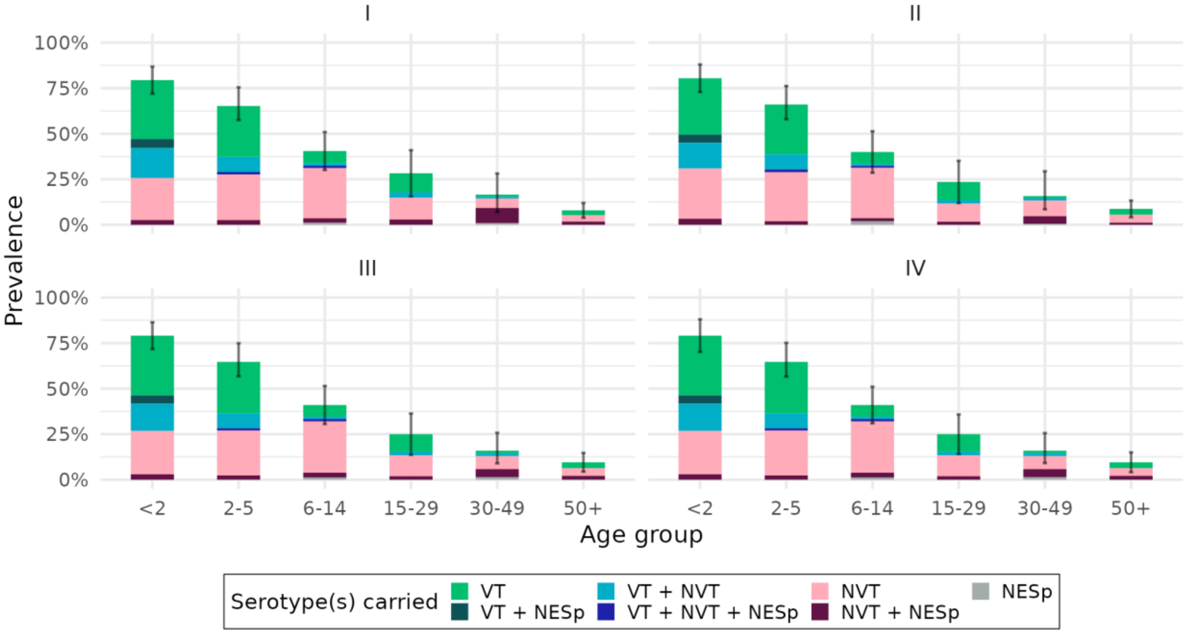

**SUPPLEMENTAL FIGURE C4. PREVALENCE AND SEROTYPE DISTRIBUTION BY AGE USING DIFFERENT WEIGHTS.**
Facets show the serotype distribution weighted by I) Age and gender, II) Age and household size, III) Age only, and IV) unweighted. Bars show the estimated prevalence of pneumococcal serotypes by age group, weighted for age and gender. Error bars show 95% confidence intervals around overall pneumococcal carriage prevalence. Colours show the prevalence of serotypes that are carried; VT: only vaccine type(s), NVT: only non-vaccine type(s); NT: only non-encapsulated type(s); and where applicable VT + NVT: both vaccine- and non-vaccine type(s). Multiple carriage with non-encapsulated type(s) is shown as a darker shading.

*Carriage prevalence compared to other settings*

We compared overall carriage prevalence and VT and NVT carriage prevalence by age with that observed in Kilifi County in Kenya, Brikama Local Government Area in the Gambia, Karonga District in Malawi, and Sheema North sub-district in Uganda (4–7). Overall carriage prevalence by age is very similar to carriage prevalence in Kenya, Malawi, and Uganda, but lower than prevalence reported in Gambia, for all age groups. A similar pattern is observed for VTs, though NVT prevalence is more similar to Malawi and Uganda than for Kenya, where it was higher. Note microbiological techniques and VT definitions (as PNEUMOSIL carriage in our Digaale dataset, as Prevenar-13 carriage in the Malawi dataset, as PCV7 carriage in the Gambian dataset, and as Synflorix carriage in the Kenyan and Ugandan datasets) differed between studies, which limits comparability.

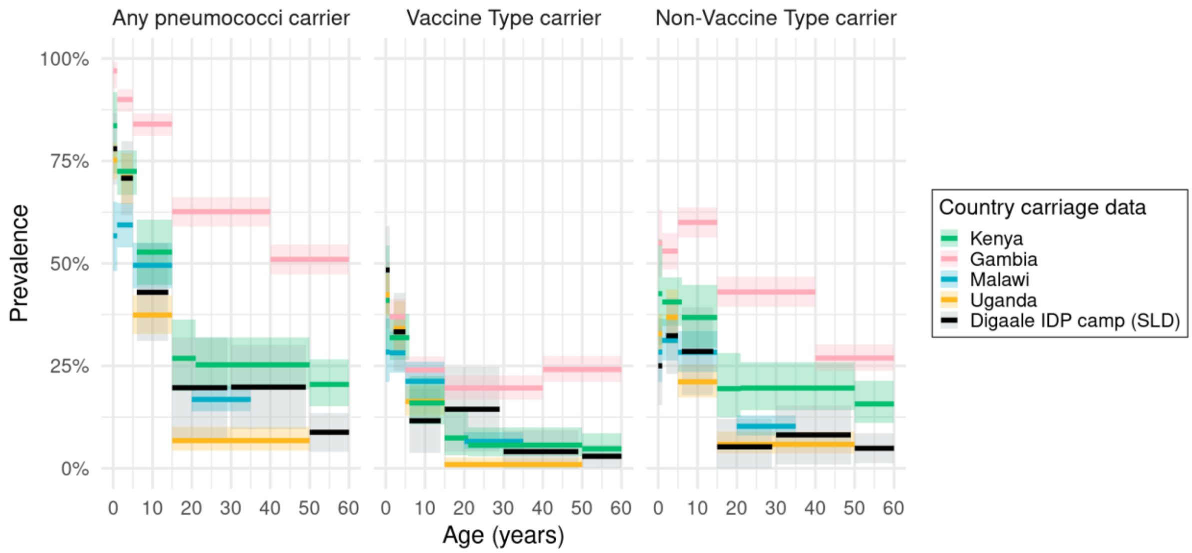

**SUPPLEMENTAL FIGURE C5. PREVALENCE BY AGE COMPARED TO DIFFERENT SETTINGS.** Pneumococcal prevalence by age in Digaale IDP camp (black), compared to prevalence observed in Kenya (green), the Gambia (pink), Malawi (blue), and Uganda (yellow). Prevenar 13 serotypes are used to define VTs. Thick lines show the age-specific carriage prevalence, and shaded areas their associated 95% binomial confidence interval. Post-stratification weights on age and gender are applied to the Digaale estimates. Studies used different age categorisations: these are shown as the width of each estimate. Facet columns show overall pneumococcal carriage prevalence, prevalence of vaccine types only, and prevalence of non-vaccine types.

*Contribution of different age groups towards the age-specific exposure to pneumococcus*

We estimated the contribution of different age groups towards the age-specific exposure to pneumococcus by combining the estimated contact matrices (8) with prevalence estimates by age (Figure 4 in main manuscript). We assessed the uncertainty around these estimates by taking 1,000 bootstrap samples of the dataset to calculate carriage prevalence and contact matrices, and calculated bootstrapped confidence interval as the 2.5% and 97.5% quantiles of estimated values over all datasets (Supplemental Table C6).

**SUPPLEMENTAL TABLE C6. The contribution of different age groups towards the age-specific exposure to pneumococcus.**

| **Contactee age group** | **Contactor age group** | | | | | |
|---|---|---|---|---|---|---|
| | *<2* | *2-5* | *6-14* | *15-29* | *30-49* | *50+* |
| *<2* | 7.6% (4.2 - 12.1) | 10.2% (8.0 - 12.9) | 5.5% (3.9 - 7.8) | 7.7% (5.4 - 10.9) | 10.0% (7.6 - 12.9) | 6.3% (4.3 - 8.9) |
| *2-5* | 39.3% (31.7 - 47.9) | 44.8% (37.5 - 52.6) | 26.0% (19.9 - 32.1) | 14.6% (10.5 - 19.9) | 21.7% (16.8 - 27.9) | 19.8% (14.6 - 25.8) |
| *6-14* | 24.6% (17.6 - 33.0) | 29.8% (22.4 - 37.6) | 51.0% (41.8 - 60.1) | 22.5% (15.2 - 30.7) | 23.7% (16.7 - 30.6) | 25.7% (17.9 - 35.0) |
| *15-29* | 14.3% (7.7 - 21.3) | 6.9% (3.6 - 10.8) | 9.3% (4.8 - 14.4) | 39.9% (24.1 - 53.0) | 19.2% (10.3 - 27.7) | 18.2% (9.6 - 26.8) |
| *30-49* | 11.9% (5.5 - 19.1) | 6.6% (3.1 - 10.9) | 6.3% (2.9 - 10.9) | 12.4% (5.9 - 20.7) | 20.4% (10.5 - 31.0) | 20.0% (9.8 - 29.9) |
| *50+* | 1.8% (0.7 - 3.4) | 1.4% (0.6 - 2.6) | 1.6% (0.7 - 3.2) | 2.8% (1.2 - 5.2) | 4.7% (2.0 - 8.4) | 9.4% (4.0 - 16.3) |

Values denote mean and bootstrapped 95% confidence intervals over 1,000 bootstrap samples of the estimated proportion of all contacts made by a contactor of age $j$ (columns), that are with contactees that are carrying pneumococci of age $i$ (rows).

### Section D. Invasive pneumococcal disease estimates

We estimated the likely proportion of invasive pneumococcal disease in the population in Digaale covered by different PCV products by applying serotype specific estimates of invasiveness to our observed carriage estimates.

*Age- and serotype-specific invasiveness*

Løchen et al (9) estimated the progression rate from carriage to invasive disease as the number of cases per carrier per year in a meta-analysis of several *Streptococcus pneumoniae* datasets using their *progressionEstimation* RStan package. They estimated progression rates separately for children and adults.

We took their reported median and 95% credible intervals values from invasiveness estimates for serotypes in children and adults, and fitted lognormal distributions to each set of values in order to recover their posterior distributions.

For each serotype *s*, in each dataset *a*, we assume that the invasiveness values can be described by a lognormal distribution with mean $\mu_{s,a}$ and standard deviation $\sigma_{s,a}$ (on the log-scale).

$$v_{s,a} \sim \text{lognormal}(\mu_{s,a}, \sigma_{s,a})$$

As the median value of any lognormal distribution is equivalent to $e^{\mu}$, we parameterized the log-mean for $v_{s,a}$ as $\mu_{s,a} = \log y_{s,a,0.5}$, where $y_{s,a,0.5}$ is the reported median estimate for serotype *s* in dataset *a.* We then optimized the value for $\sigma_{s,a}$ using the *optimize* function in base *R* to minimize $g_{(\sigma_{s,a})}$: the log-least squares of the difference between the reported boundaries of the 95% credible interval for $v_{s,a}$, $y_{s,a,0.025}$ and $y_{s,a,0.975}$, and corresponding quantile distribution for the lognormal distribution evaluated at 0.025 and 0.975. We took the log of the values to minimize the large difference in scale between the lower and upper boundary value.

$$g(\sigma_{s,a}) = \log\left(\left(y_{s,a,0.025} - Q_{s,a}(0.025)\right)^2 + \left(y_{s,a,0.975} - Q_{s,a}(0.975)\right)^2\right)$$

Where $Q_{s,a}(p)$ is the quantile distribution of the lognormal distribution used to describe $v_{s,a}$ evaluated at *p*.

Supplemental Figure D1 shows the fitted lognormal distributions against the reported values estimated by Løchen et al. The 95% quantile values of the fitted distributions were in broad agreement with the reported 95% credible intervals, with only some minor discrepancies at primarily the lower tails of some distributions.

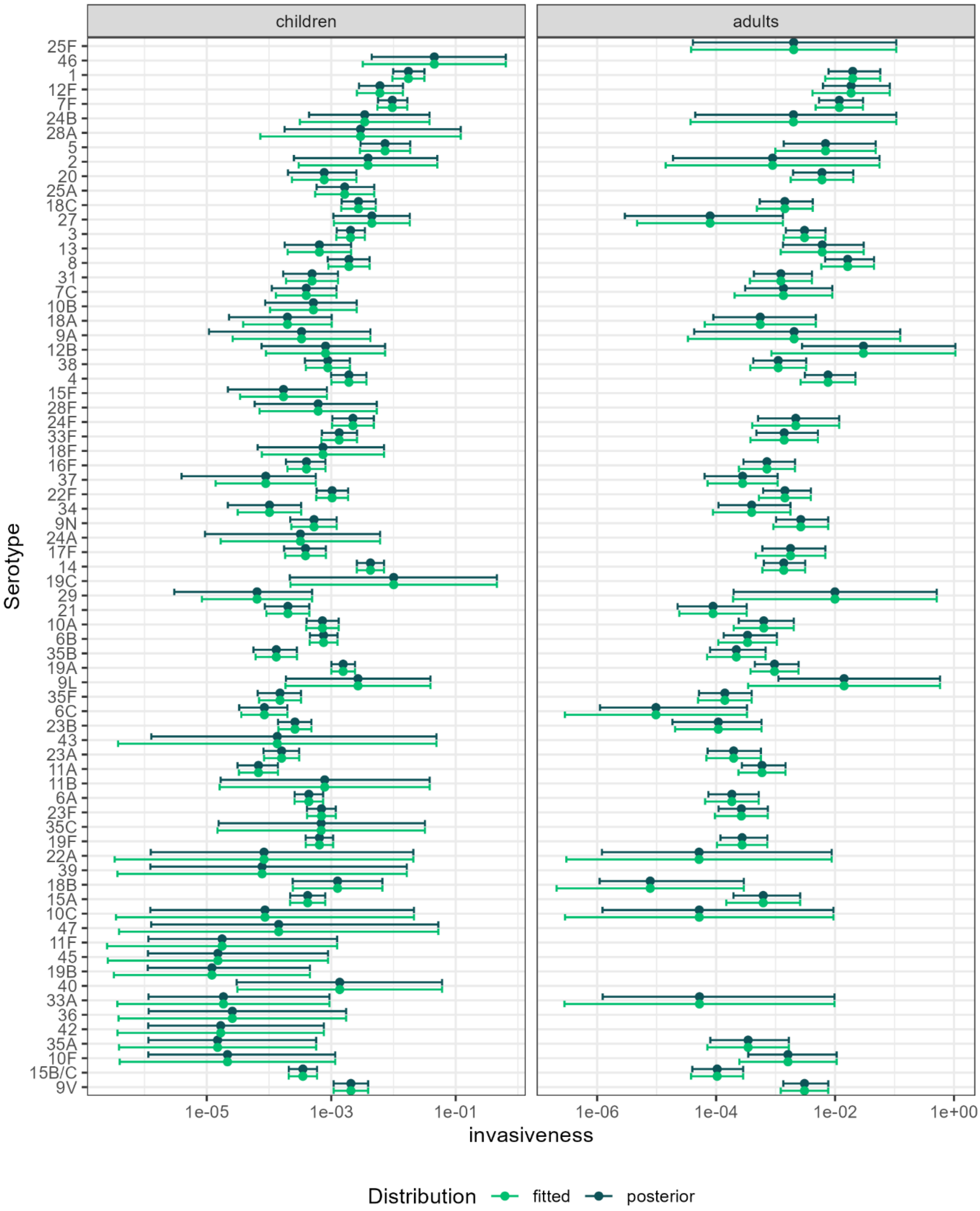

**SUPPLEMENTAL FIGURE D1. AGE AND SEROTYPE SPECIFIC INVASIVENESS VALUES.** Dark-green values show the median and 95% credible intervals reported by Løchen et al (9). Light-green values show the median and 95% quantile values from the refitted lognormal distributions.

*Estimating invasiveness in Digaale*

We applied the invasiveness estimates to our carriage estimates by generating 10,000 bootstrap samples from our dataset where we resampled participants with replacement. In each bootstrap sample, we sampled age- and serotype-specific invasiveness values from the fitted invasiveness distributions, and applied these values to carriers of that serotype in the bootstrapped dataset. Poststratification weights were recalculated in each bootstrap dataset to ensure representativity of population-level estimates.

We assumed the same invasiveness value for any carried pneumococci of the same serotype, regardless of their abundance or the number of other serotypes carried by an individual. We applied invasiveness values sampled from invasiveness distributions for children to serotypes carried by individuals aged <18y, and values from distributions for adults to all other serotypes. We ignored any invasive disease caused by non-encapsulated serotypes, by assuming an invasiveness value of 0.

The Løchen et al dataset provided invasiveness values for 33/34 encapsulated serotypes identified in children, and for 17/20 encapsulated serotypes identified in adults. When available, we replaced any missing values by the average invasiveness of serotypes in the same age and serogroup. Values for serotype 20B in both children and adults were replaced with age-specific estimated values for serotype 20. Values for serotype 19B in adults were replaced by the mean values for serotypes 19A and 19F. Values for serotype 41A in adults were replaced by the median of all adult invasiveness values, as no estimate was available for any serotype in serogroup 41.

The total number of IPD cases expected within one year in bootstrap sample *i* excluding those in PCV product *p* was calculated as:

$$d_{i,p} = \sum_{x=1}^{N} \left( w_{i,x} \sum_{s=1}^{S} v_{i,s,a} I_x(s) \left(1 - V_p(s)\right) \right)$$

Where *N* is the total number of individuals in the dataset, $w_{i,x}$ is the post-stratification weight calculated for individual *x* in bootstrap sample *i*, *S* is the total number of unique serotypes identified in Digaale, $v_{i,s,a}$is the invasiveness value sampled for serotype s in bootstrap sample i, for individuals of age *a*, *a* is the index of the age-group for individual x (<18y or ≥18y), $I_x(s)$ is an indicator function that returns 1 if individual x carries serotype s, and 0 otherwise, and $V_p(s)$ is an indicator function that returns 0 if serotype *s* is not included in PCV product *p*, and 1 if it is included. $V_0$ denotes no vaccination, and returns 0 for all serotypes *s*. for serotype s and individual x, which returns 1 if individual estimate for serotype.

For each vaccine PNEUMOSIL, Synflorix, Prevenar 13, Vaxneuvance, and Prevenar 20, we calculated the proportion of the total invasiveness i) unweighted in the sampled dataset and ii) at the population level by applying post-stratification weights in all ages, those <5y, and those in age groups <2y, 2-5y, 6-14y, 15-29y, 30-49y, and 50+y.

The proportion of total invasiveness caused by serotypes covered by vaccine product *p* in bootstrap sample *i* was then calculated as $1 - \frac{d_{i,p}}{d_{i,0}}$

We reported the median and 95% quantile values of that proportion across all bootstrap samples.

*Results*

We estimate that between 52% (95%CI 30% – 65%) of all IPD cases were caused by serotypes included in the PNEUMOSIL vaccine. The estimated proportion for serotypes covered by Synflorix serotypes was slightly higher (61%; 43 – 76), with higher proportions for serotypes included in higher valency vaccines (Prevenar 13: 75%, 60 – 85; Vaxneuvance: 76%, 61 – 86; and Prevenar 20: 82%, 68 – 89) (Supplemental Figure D2 and Supplemental Table D1). While the proportion tended to be the lowest for serotypes included in the

PNEUMOSIL vaccine and the highest for serotypes included in the Prevenar 20 vaccine in all age groups, this difference was especially apparent in estimated IPD in people aged 15y and older.

These results indicate that a substantial proportion of current IPD is likely caused by serotypes included in any of the PCVs. We caution against overinterpretation of these results, as no data is available to validate these estimates, and uncertainty around the estimates is wide. These estimates are not estimating the potential impact of PCVs, which depend on a multitude of factors including their vaccine coverage, serotype-specific vaccine efficacy, immunological cross-reactivity with uncovered serotypes, and indirect effects including serotype replacement of VTs with NVTs.

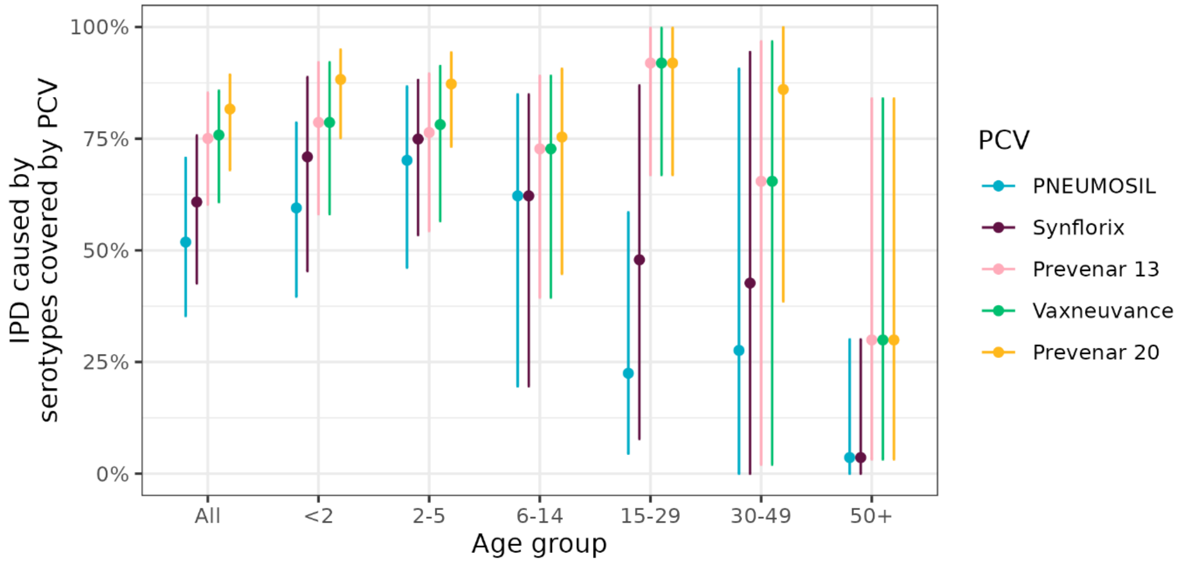

**SUPPLEMENTAL FIGURE D2. ESTIMATED PROPORTION OF IPD CASES CAUSED BY SEROTYPES COVERED BY PCVS.** Points shows the median and lines the 95% uncertainty interval estimated from 10,000 bootstrapped samples, for different PCVs, by age group.

**TABLE D1. Proportion of current IPD covered by PCVs**

| Age group | PNEUMOSIL[a] | Synflorix[b] | Prevenar 13[c] | Vaxneuvance[d] | Prevenar 20[e] |
|---|---|---|---|---|---|
| *All* | 51.9% (35.3 - 70.7) | 60.8% (42.6 - 75.7) | 75.1% (60.3 - 85.3) | 75.8% (60.8 - 85.7) | 81.6% (68.0 - 89.3) |
| *<2* | 59.5% (39.7 - 78.6) | 70.9% (45.4 - 88.8) | 78.6% (58.2 - 92.1) | 78.6% (58.2 - 92.1) | 88.3% (75.2 - 94.9) |
| *2-5* | 70.2% (46.1 - 86.7) | 74.9% (53.4 - 88.1) | 76.4% (54.4 - 89.6) | 78.2% (56.6 - 91.2) | 87.3% (73.2 - 94.3) |
| *6-14* | 62.2% (19.6 - 84.9) | 62.2% (19.6 - 84.9) | 72.7% (39.5 - 89.0) | 72.7% (39.5 - 89.0) | 75.4% (44.8 - 90.6) |
| *15-29* | 22.5% (4.5 - 58.5) | 47.9% (7.8 - 86.9) | 91.9% (66.9 - 99.8) | 91.9% (66.9 - 99.8) | 91.9% (66.9 - 99.8) |
| *30-49* | 27.6% (0.0 - 90.6) | 42.7% (0.0 - 94.3) | 65.5% (2.1 - 96.7) | 65.5% (2.1 - 96.7) | 86.0% (38.6 - 99.9) |

| | | | | | |
|---|---|---|---|---|---|
| *50+* | 3.6% (0.0 - 30.0) | 3.6% (0.0 - 30.0) | 29.9% (3.2 - 84.0) | 29.9% (3.2 - 84.0) | 29.9% (3.2 - 84.0) |

Median estimate and 95% quantile values of proportion of current IPD cases that are caused by serotypes covered by PCVs, across 10,000 bootstrap samples of participant datasets and age-and serotype specific invasiveness estimates.

a. Serotypes 1, 5, 6A, 6B, 7F, 9V, 14, 19A, 19F, and 23F.
b. Serotypes 1, 4, 5, 6B, 7F, 9V, 14, 18C, 19F, and 23F.
c. Serotypes 1, 3, 4, 5, 6A, 6B, 7F, 9V, 14, 18C, 19A, 19F, and 23F.
d. Serotypes 1, 3, 4, 5, 6A, 6B, 7F, 9V, 14, 18C, 19A, 19F, 22F, 23F, and 33F.
e. Serotypes 1, 3, 4, 5, 6A, 6B, 7F, 8, 9V, 10A, 11A, 12F, 14, 15B, 18C, 19A, 19F, 22F, 23F, and 33F.

### References used in Supplemental Material

1. Lex A, Gehlenborg N, Strobelt H, Vuillemot R, Pfister H. UpSet: Visualization of Intersecting Sets. IEEE Transactions on Visualization and Computer Graphics. 2014 Dec;20(12):1983–92.

2. Pell CL, Ortika BD, Van Zandvoort K, Wee-Hee A, Mulholland EK, Checchi F, et al. Effect of transport conditions on recovery of *Streptococcus pneumoniae*. Abstract presented at: The 12th International Symposium on Pneumococci and Pneumococcal Diseases (ISPPD); 2022 Jun; Toronto, Canada.

3. Lumley T. survey: analysis of complex survey samples. 2020.
